# Mathematical Model of a Personalized Neoantigen Cancer Vaccine and the Human Immune System: Evaluation of Efficacy

**DOI:** 10.1101/2021.01.08.21249452

**Authors:** Marisabel Rodriguez Messan, Osman N. Yogurtcu, Joseph R. McGill, Ujwani Nukala, Zuben E. Sauna, Hong Yang

## Abstract

Cancer vaccines are an important component of the cancer immunotherapy toolkit enhancing immune response to malignant cells by activating CD4^+^ and CD8^+^ T cells. Multiple successful clinical applications of cancer vaccines have shown good safety and efficacy. Despite the notable progress, significant challenges remain in obtaining consistent immune responses across heterogeneous patient populations, as well as various cancers. We present as a proof of concept a mechanistic mathematical model describing key interactions of a personalized neoantigen cancer vaccine with an individual patient’s immune system. Specifically, the model considers the vaccine concentration of tumor-specific antigen peptides and adjuvant, the patient’s major histocompatibility complexes I and II copy numbers, tumor size, T cells, and antigen presenting cells. We parametrized the model using patient-specific data from a recent clinical study in which individualized cancer vaccines were used to treat six melanoma patients. Model simulations predicted both immune responses, represented by T cell counts, to the vaccine as well as clinical outcome (determined as change of tumor size). These kinds of models have the potential to lay the foundation for generating *in silico* clinical trial data and aid the development and efficacy assessment of personalized cancer vaccines.

**Author summary:** Personalized cancer vaccines have gained attention in recent years due to the advances in sequencing techniques that have facilitated the identification of multiple tumor-specific mutations. This type of individualized immunotherapy has the potential to be specific, efficacious, and safe since it induces an immune response to protein targets not found on normal cells. This work focuses on understanding and analyzing important mechanisms involved in the activity of personalized cancer vaccines using a mechanistic mathematical model. This model describes the interactions of a personalized neoantigen peptide cancer vaccine, the human immune system and tumor cells operating at the molecular and cellular level. The molecular level captures the processing and presentation of neoantigens by dendritic cells to the T cells using cell surface proteins. The cellular level describes the differentiation of dendritic cells due to peptides and adjuvant concentrations in the vaccine, activation, and proliferation of T cells in response to treatment, and tumor growth. The model captures immune response behavior to a vaccine associated with patient specific factors (e.g., different initial tumor burdens). Our model serves as a proof of concept displaying its utility in clinical outcomes prediction, lays foundation for developing *in silico* clinical trials, and aids in the efficacy assessment of personalized vaccines.

## Introduction

Cancer vaccines, alone or in conjunction with other immune modulators (e.g., checkpoint inhibitors) are among the most promising therapeutic options for many human cancers [1, 2]. This therapy employs either dendritic cells, T cells, DNA, RNA, viral vectors, proteins, peptides, or tumor cell lysate. The goal is exploiting the patient’s immune system to target antigens expressed only on tumor cells, in order to selectively eliminate cancer cells [1, 3–5]. Therapeutic strategies that incorporate personalized cancer vaccines have gained attention due to successes in targeting multiple tumor-specific mutations [6]. However, tumor gene expression and mutations are immensely diverse, and how they affect outcomes for individual patients remains poorly understood [7]. Tumor-specific antigens (TSAs), or neoantigens, carry the amino acid substitutions derived from random somatic mutations that are expressed only on tumor cell surface. These mutations are highly patient- and tumor-specific [2, 6, 8, 9].

With the advent of accurate and inexpensive Next Generation Sequencing techniques, routine sequencing of the DNA and RNA from cancer cells has become possible. Subsequent workflows using bioinformatic tools allow the identification of neo-sequences. These neo-sequences carry somatic mutations exhibiting sequences that are different from the wild type, and hence can be recognized as nonself by the host immune system, with a high probability of eliciting a cancer-specific immune response. From the large number of neo-sequences identified, the selection of the few neo-sequences to be used as neoantigens in cancer vaccines largely is based on the affinity of the neoantigens peptides to the patient’s major histocompatibility complex-I (MHC-I) and MHC-II proteins [7]. Individualized immunotherapies designed based on these principles have the potential to be specific, efficacious, and safe [9–11]. Eliciting immune responses to protein targets not found on normal cells reduces the probability of immune toxicities [9].

Despite some successes with the strategy of designing personalized neoantigen cancer vaccines described above, significant challenges remain. Selection of the candidate neoantigen based on peptide-MHC binding affinity focuses on just one step of a complex immunological cascade. The peptide-MHC engagement is a necessary, but not sufficient, step for the immune response to occur. More comprehensive models are required to effectively evaluate the potential neo-sequences to select those that will be the most efficacious as peptide vaccines. These strategies must have a rapid turnaround time, as the window of opportunity to treat an advanced cancer patient is limited. Also, there is lack of qualified predictive biomarkers for determining whether the therapy will be effective. Quantitative modeling approaches provide a useful toolkit for studying the interplay between tumor cells and the immune system [12]. These approaches enable a quantitative understanding of immune response kinetics following neoantigen-based peptide vaccine treatment. Insights gained from challenges can be used to design better vaccines and evaluate the potential candidate vaccines *in silico*. The models also can guide such decisions on treatment regimens such as dosing and infusion frequencies [13].

Mathematical modeling in biology and medicine, especially oncology [14, 15], can be a powerful tool for the generation of experimental and clinical hypotheses to aid in study design [16, 17]. A mathematical model simplifies complex biological systems, provides insights into known mechanisms of the systems, and can aid researchers and clinicians better understand the disease and treatment [13, 17]. Models can facilitate in the development of testable hypotheses by providing quantitative estimates for numerous outcomes produced by dynamic interactions between various biological mechanisms. To date, several mathematical models have been developed to predict the effect of mono or combination immunotherapies against cancer [13, 18–25]. Although these studies provide insights into the interactions between treatments and immune response by modeling cell- and/or tissue-level immune dynamics, components of immune dynamics at the molecular level are lacking. The addition of this level of granularity to a mathematical model describing the immune and clinical response to a cancer vaccine is critical. Key molecular events that may impact the immune response and clinical outcome include, antigen processing and presentation, and MHC-T cell binding. Thus, a model that successfully describes the immune response elicited by a cancer vaccine (and the subsequent effect on tumors) will need to include the molecular parameters and events that drive the immunological cascade. The model presented here addresses this critical unmet need.

## Results

### Model parametrization for individual patients based on their T cell response

Our mathematical model is designed to quantify the effect of a personalized peptide cancer vaccine, which is a mixture of peptide neoantigens with an adjuvant, on (1) T cell response and (2) change in tumor size. To parametrize our model, we fitted the six individual patients’ longitudinal T cell response data extracted from a published study [26] as described in the Supplementary S1 Appendix to obtain patient-specific initial conditions and model parameters, i.e., an individual patient’s T cell response to peptides, initial tumor cell count, HLA alleles, and the peptide amino acid sequences of the personalized vaccine. Patients’ activated T cell counts data over time and the model fits with the highest adjusted *R*^2^ are shown in Fig 1. The patient-specific best-fit parameter values are tabulated in Table 1. Detailed description of these parameters (labeled with ⋆) is on Supplementary S1 Table.

**Table 1.**
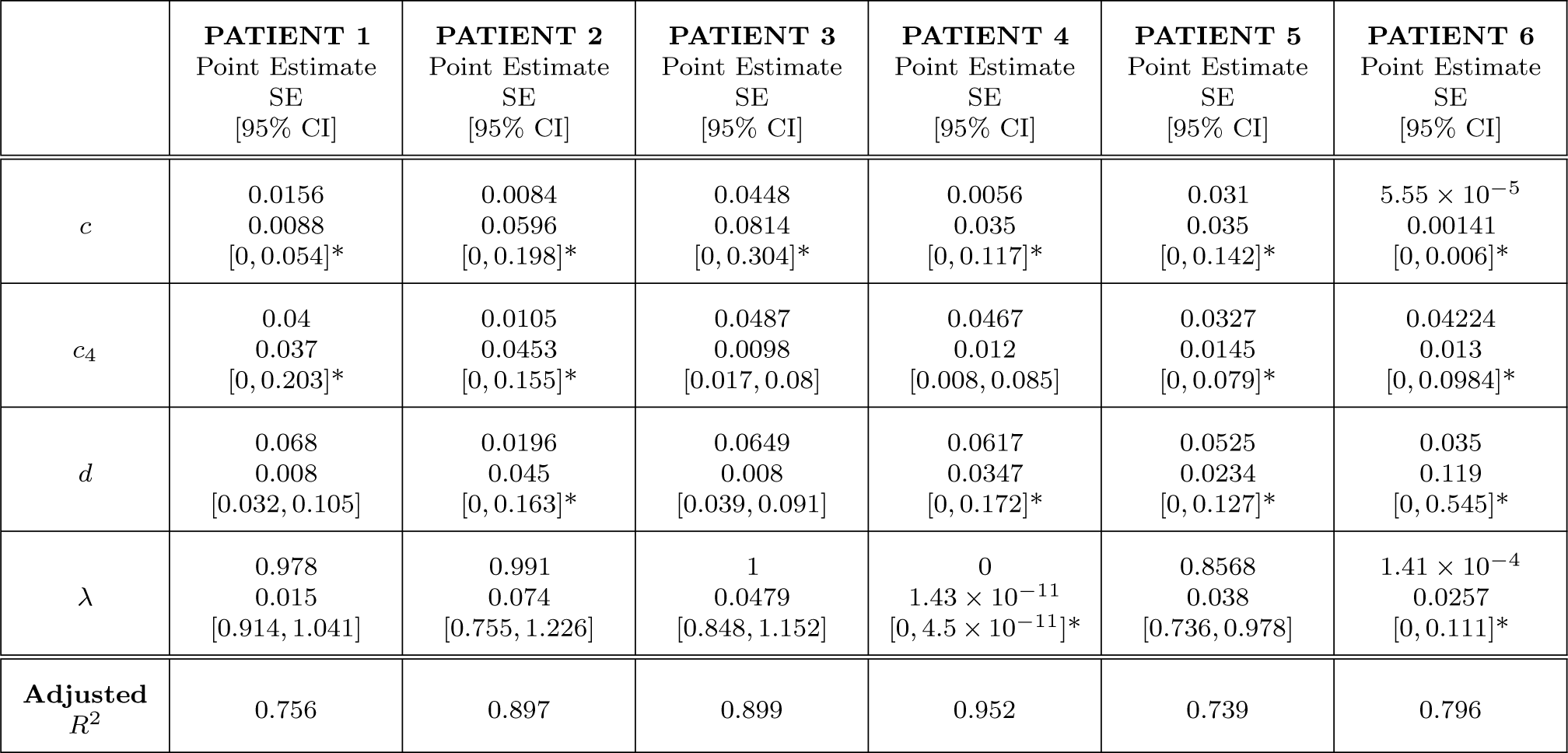
Best-fit values of model parameters for six melanoma patients in study [26]. *c*: Maximum CD8 T cells recruitment rate; *c*_4_: Maximum CD4^+^ T cell production rate; *d*: Maximum lysis rate by activated T cells; *λ*: dependence of lysis rate on the effector/target ratio constant. [95% CI]* denotes modified confidence interval by replacing negative lower endpoint with zero as described in [27].

**Fig 1.**
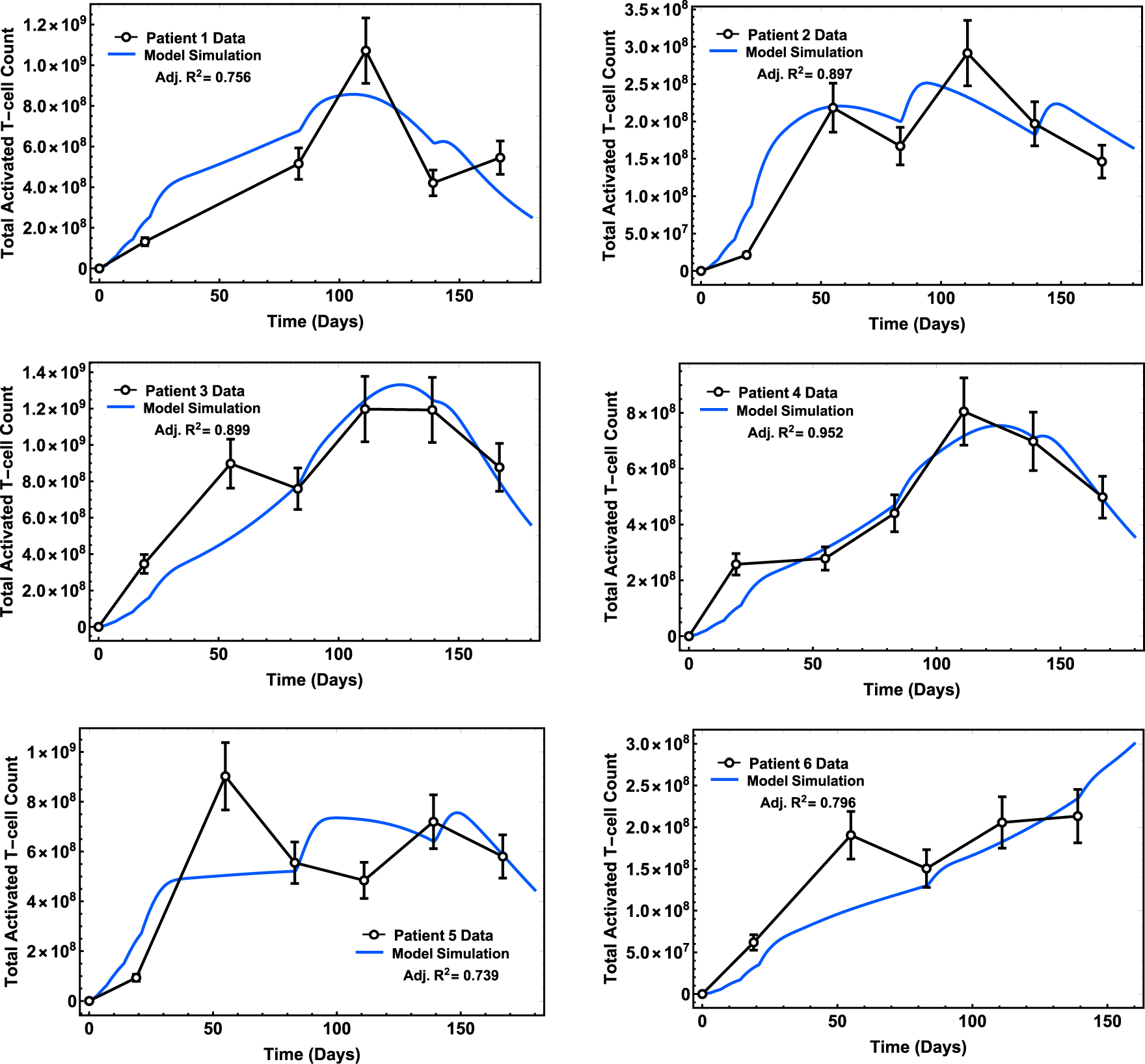
Time profiles of T cell response of patients. Model fits with the highest adjusted *R*^2^ are plotted alongside clinical data from a Phase 1 study [26] (open circles with error bars, 15% standard error measurement; see S1 Appendix for details; blue arrows indicate days when prime and booster vaccine doses were administered).

### Longitudinal model forecasts of change of tumor size in clinical trial patients

To further explore the utility of our model, we used the best-fit parameter values in Table 1 to predict the change of tumor size of individual patients. The clinical trial [26], briefly reported that four patients (1, 3, 4, and 5), initially diagnosed with Stage 3 melanoma, exhibited no recurrence of the disease following the treatment regimen with the cancer vaccine. On the other hand, patients 2 and 6, initially diagnosed with Stage 4 melanoma, showed disease recurrence after the vaccine treatment. Using each patient’s best-fit parameter values (Table 1), we simulated the tumor cell dynamics under influence of the vaccine. We computationally estimated the tumor size (diameter in millimeters; cell counts highlighted in yellow) 200 days after treatment initiation for all six patients (see Fig 2). For patients 2 and 6, we further compared model-predicted tumor size at days 196 and 146, respectively, with CT scan images and sizes obtained in the study [26] at these two time points, respectively.

**Fig 2.**
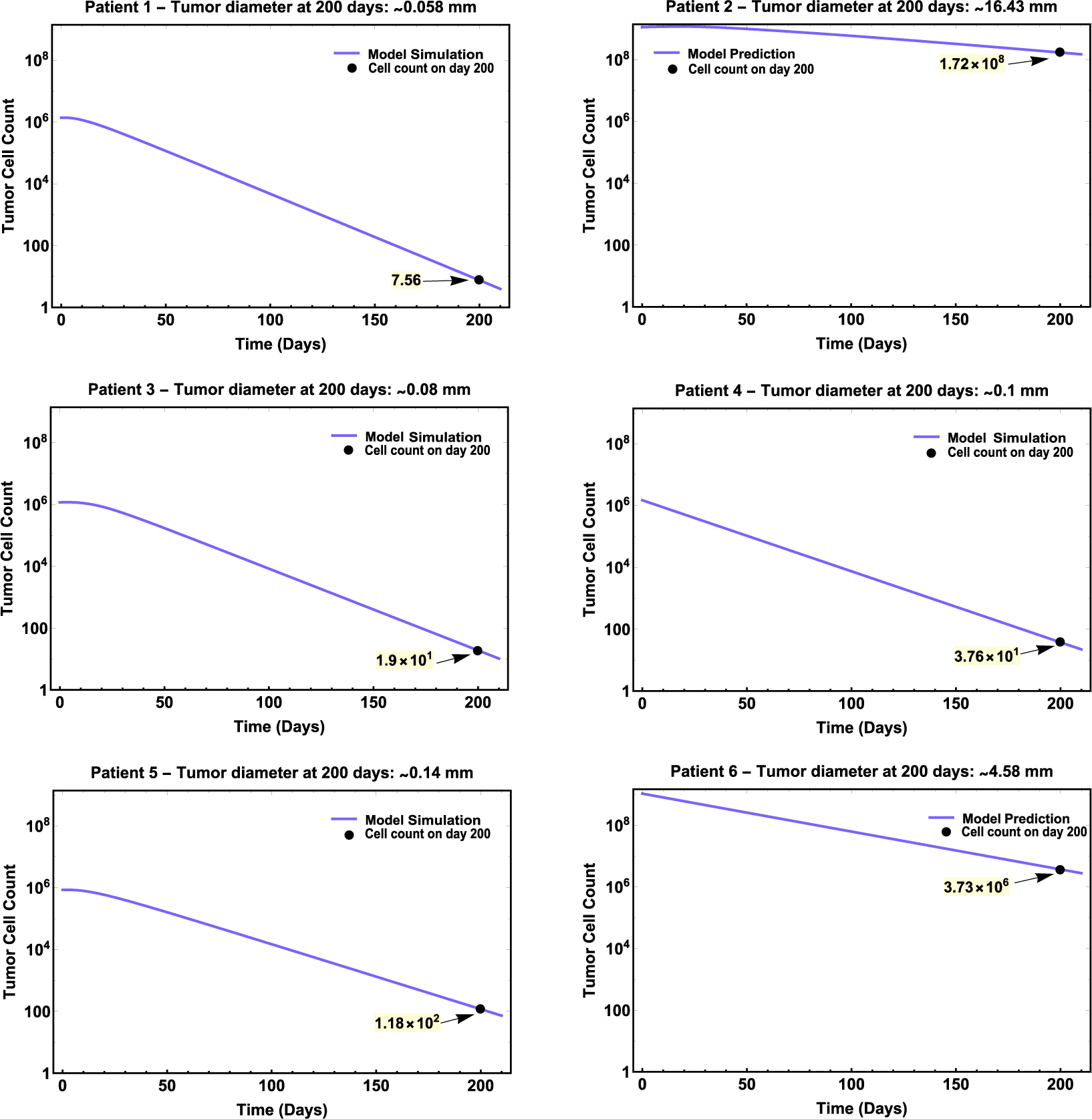
Longitudinal tumor burden predictions for patients 1-6 with quantitative tumor diameter predictions on day 200. Model simulations are plotted in log-scale with respect to tumor cell count. Model-predicted malignant cell count on specified day (black dot) is shown highlighted in yellow and tumor size (diameter) was estimated in millimeters from tumor cell count.

Model predictions of tumor size for patient 1, 3, 4, and 5 (no reported disease recurrent) showed a relatively greater decrease of tumor size (or cell count) with respect to patients 2 and 6 (with report of disease recurrent) at the end of vaccine treatment. Our model predictions on residual malignancies are qualitatively consistent with reported clinical are consistent of non-recurrence versus recurrence (see Fig 2).

The CT scans for Patient 2 were obtained 8 weeks after last booster vaccine (day 196) showing a tumor measuring approximately 21 mm, and our model predicted a tumor diameter of approximately 16.7 mm in diameter (Fig 3 - left).

**Fig 3.**
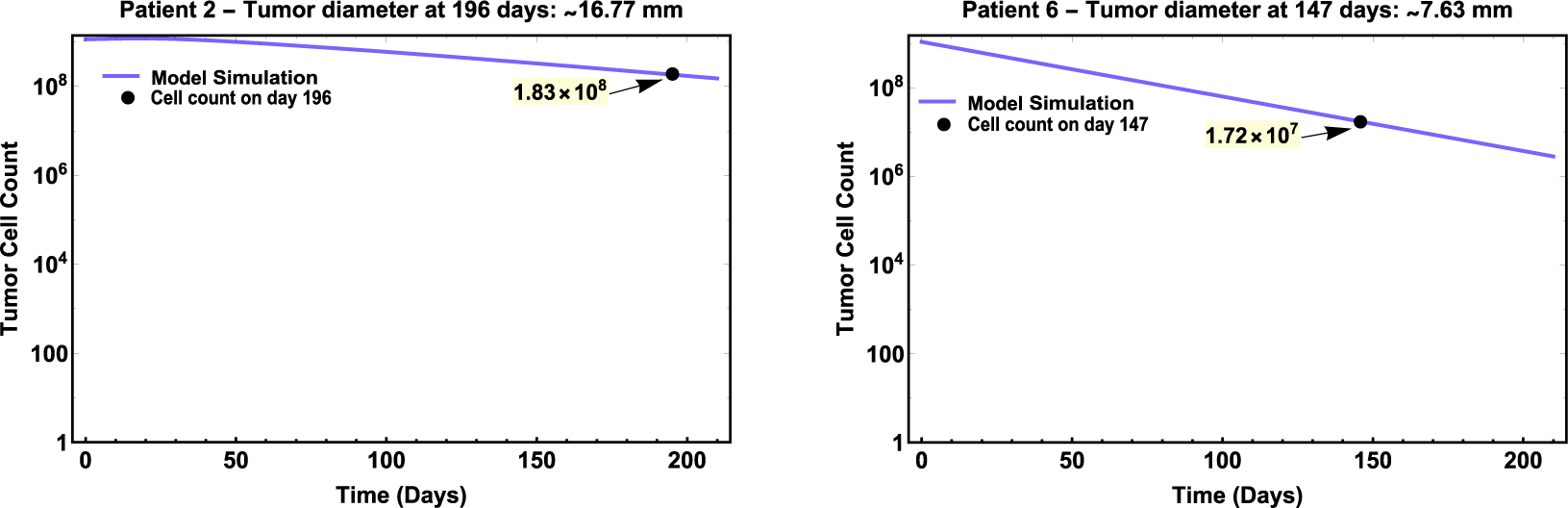
Longitudinal tumor burden predictions Patients 2 and 6 with quantitative tumor diameter predictions on day 196 and 147, respectively. Model simulations are plotted in log-scale with respect to tumor cell count. Model-predicted malignant cell count on specified day (black dot) is shown highlighted in yellow and tumor size (diameter) was estimated in millimeters from tumor cell count.

**Fig 4.**
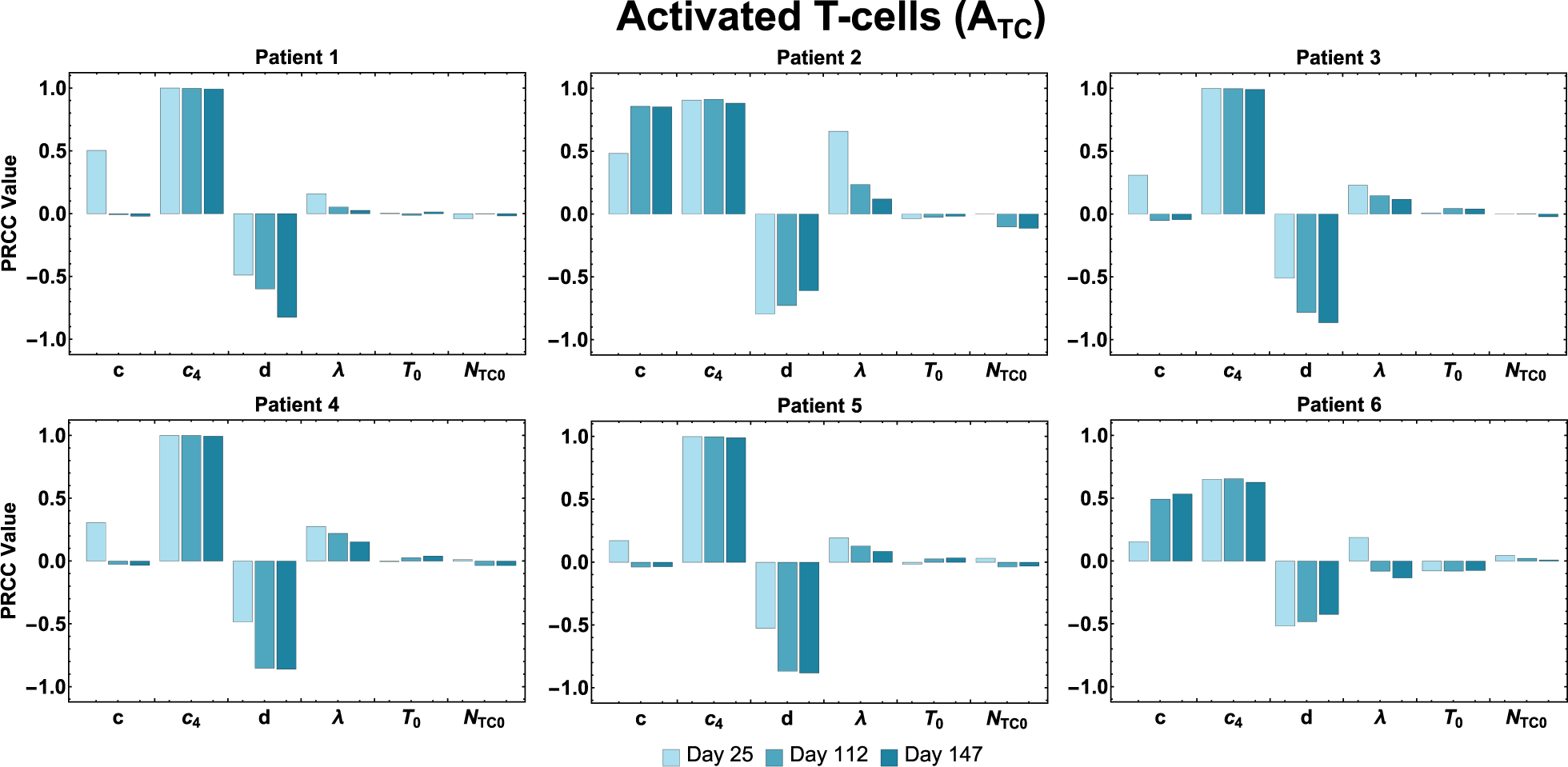
Activated T cell population sensitivity. PRCC values are shown for the estimated parameters using the total activated T cell population as the output of interest.

**Fig 5.**
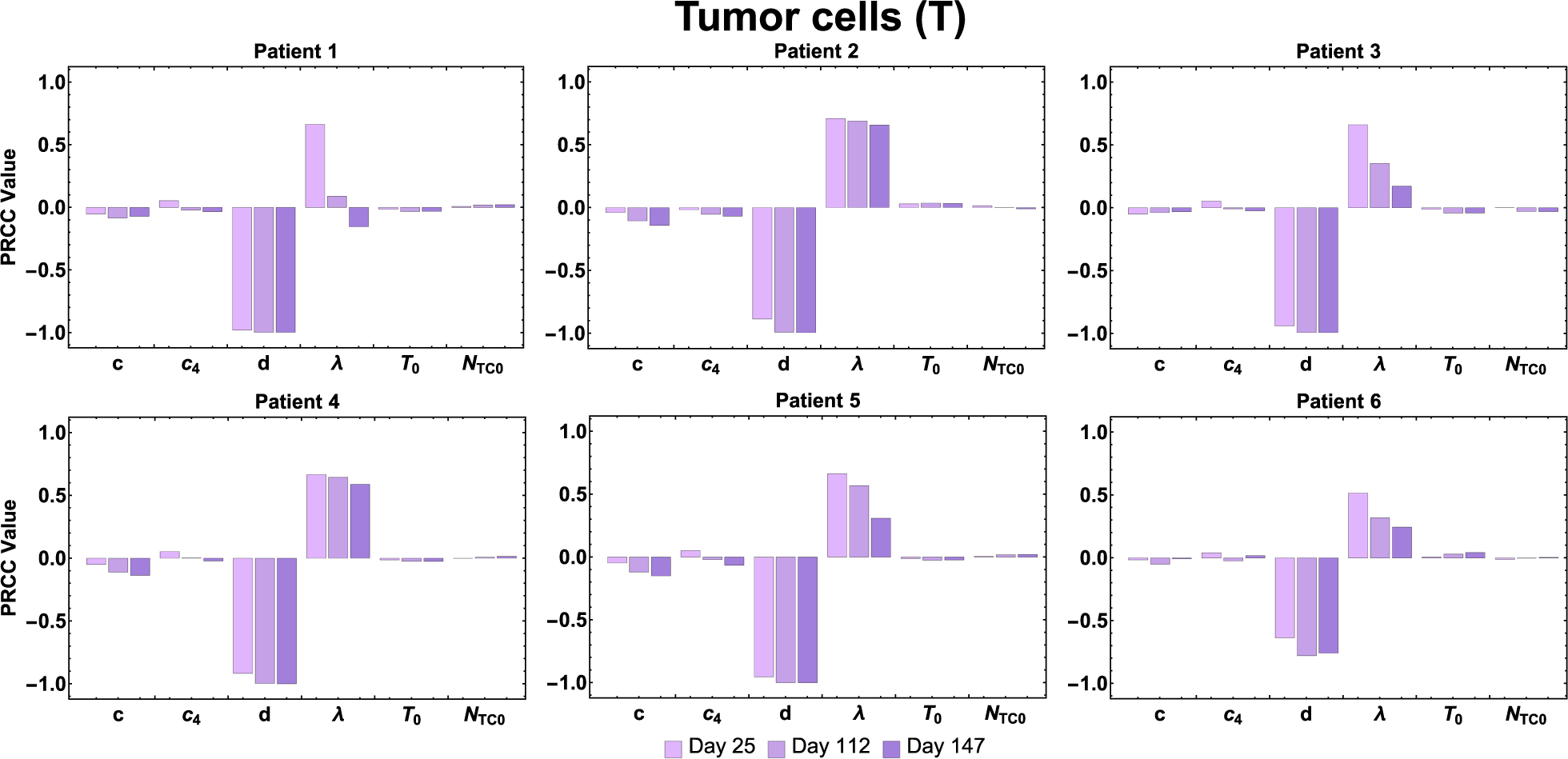
Tumor cell population sensitivity. PRCC values are shown for the estimated parameters using the tumor cell population as the output of interest.

For Patient 6, CT scans were obtained one week after last booster vaccine, that is on day 147 (assuming vaccine was administered on day 140 corresponding to week 20 of vaccination protocol). The CT scans showed multiple soft-tissue nodules; one of them measuring 21*×*18 mm. Our model predicted a tumor diameter size of approximately 7.63 mm in diameter for this patient (Fig 3 -right).

## Sensitivity analysis

To explore the behavior of this complex mathematical model, we performed a global sensitivity analysis. Uncertainty of model predictions is often a result of errors in the model structure compared to the real biological system, estimations of parameters using different data sets, parameter values obtained from different sources, unknown errors in the input data, and errors in model solution algorithms. Using sensitivity analysis, we can quantify how a change in the value of an input parameter changes the value of the outcome variable. The sensitivity analysis thus allows the identification of parameters that make major contributions to the uncertainty of the outcomes [28].

For all patients, the sensitivity of the outcome variable *A*_*TC*_ has shown to be consistently high throughout three time points (days 25, 112, 147) to the parameter *c*_4_ (maximum CD4^+^ T cell production rate stimulated by the interactions with tumor cells), since |*PRCC* (*A*_*TC*_, *c*_4_)| *≈* 1, i.e., magnitude of the PRCC value of parameter *c*_4_ with respect to the output of interest *A*_*TC*_ is close to 1. Also, for all patients, the variable *A*_*TC*_ was shown to be sensitive to the parameter *d* (maximum killing rate by activated CD8^+^ T cells), i.e., |*PRCC* (*A*_*TC*_, *d*)| *≈* 1. However, the sensitivity to parameter *d* for Patient 1, 3, 4, and 5 was shown to increase over time, the opposite occurred for Patient 2 and 6. For individual patients 1, 2, and 6, the variable *A*_*TC*_ was shown to be sensitive to the parameter *c* (maximum CD8^+^ T cells recruitment rate stimulated by interactions with tumor cells), in which sensitivity to *A*_*TC*_ to *c* increased over time for Patient 2 and 6.

For the second output of interest, tumor cells (*T*), PRCC values show that *d* (maximum tumor killing rate by activated CD8^+^ T cells) is consistently the most influential parameter for all patients and throughout each time point. The second most influential parameter to the sensitivity of tumor cell counts for all patients at the ‘priming phase’ (day 25) is *λ* (dependence of killing rate on the effector/target ratio constant). For Patients 2 and 4, the variable for tumor cells remains sensitive to parameter *λ* at different PRCC values throughout the rest of the phases (‘peak T cell response’ and ‘end of treatment’).

However, this analysis revealed both variables of interest, activated T cells and tumor cells (*A*_*TC*_ and *T*) were not sensitive, to initial counts of tumor cells (*T*_0_) and naïve T cells (*N*_*TC*0_) for all the time points considered.

### Application: impact of cancer vaccine treatment on clinical outcome

To explore the impact of vaccine administration at different initial tumor sizes, we predicted tumor diameter size (or cell count) 20 days after the last vaccination was administered (day 168) as a surrogate marker for clinical outcome. We selected four critical tumor diameters based on total cell counts at time of vaccination and made predictions using individual patient data and settings. Methodological details for generating Fig 6 can be found in the Supplementary S1 Appendix.

**Fig 6.**
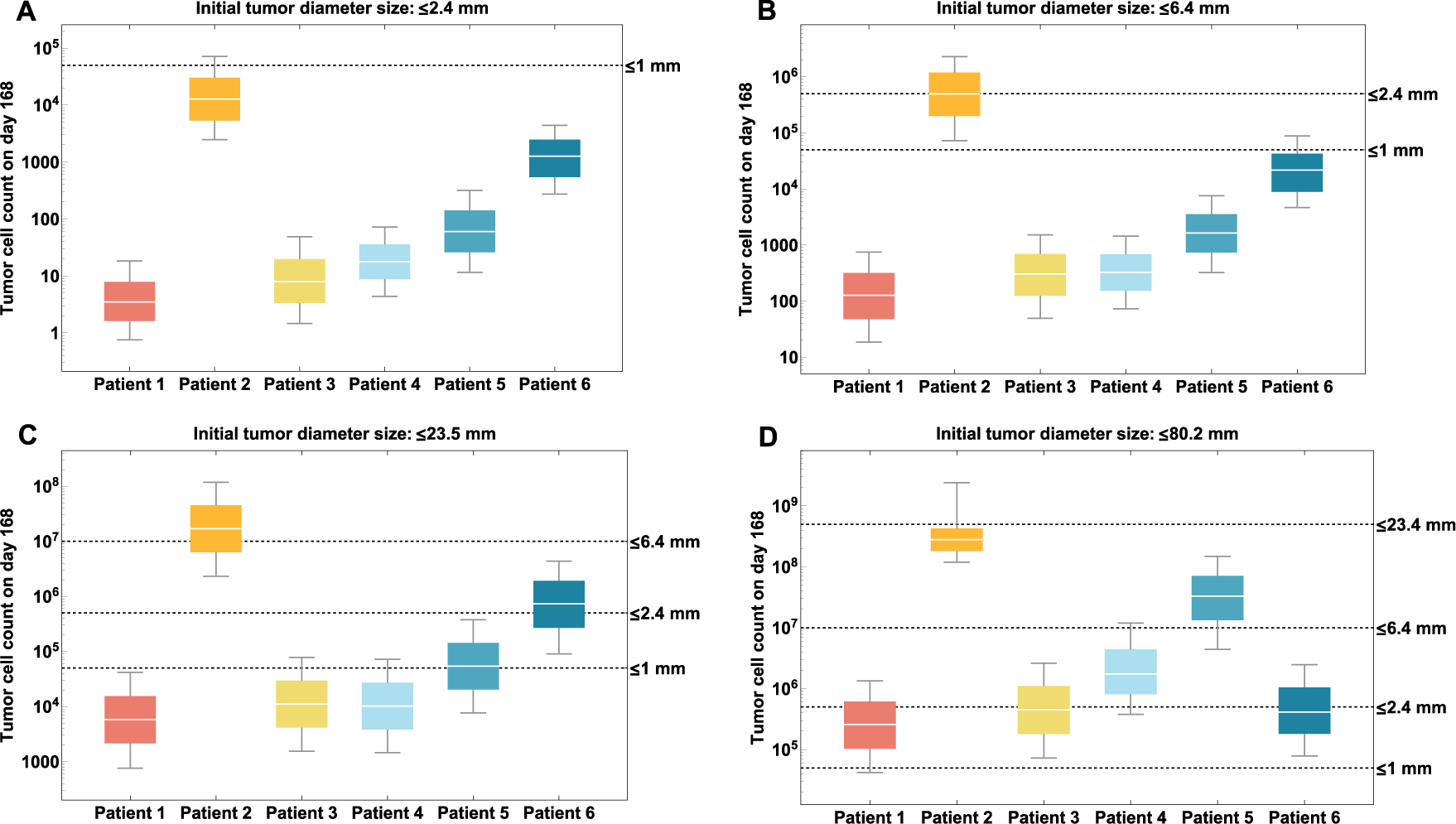
Prediction of tumor diameter by patient according to different initial tumor size at time of clinical trial enrollment. (a) Initial tumor diameter is less than or equal to 2.4 mm; (b) Initial tumor diameter is larger than 2.4 mm but smaller or equal to 6.4 mm; (c) Initial tumor diameter is larger than 6.4 mm but smaller or equal to 23.45 mm. (d) Initial tumor diameter is larger than 23.45 mm but smaller or equal to 80.2 mm.

In Fig 6, we can observe different outcomes for patients based on their profiles (i.e., HLA alleles, peptide-vaccine concentration, estimated patient-specific parameters). Fig 6A, for example, shows that all patients are able to achieve optimal tumor size reduction to less than 1 mm in diameter after vaccination if they start treatment when the tumor has a diameter of less than 2.4 mm. If patients start treatment when the tumor has a diameter larger than 2.4 mm but smaller than 6.4 mm, we predict (Fig 6B) that tumor sizes of patients 1, 3, 4, 5, and 6 would be smaller compared to Patient 2. In Fig 6C, Patients 1, 3, 4, and 5, starting treatment when tumor diameter is between 6.4 and 23.45 mm, are shown to achieve optimal tumor size reduction to less than 1 mm in diameter. Finally, Fig 6D, shows that patients who start vaccine treatment with a much larger tumor size (*>*23.6 mm) are not able to reach an optimal tumor size reduction at the end of treatment.

## Discussion

Cancer vaccines have the potential to enhance the immune response to target and eliminate cancer cells via activated CD4^+^ and CD8^+^ T cells. In this work, we focus on understanding and analyzing important mechanisms involved in the activity of personalized cancer vaccines using a mechanistic mathematical model. Our model simulations can capture immune response behavior to a vaccine associated with patient specific factors (e.g., different initial tumor burdens).

Our model currently includes a phenomenological response as a means to simulate elimination of tumor cells by activated T cells. This modeling approach has its limitations, since the neoantigen peptide vaccine’s secondary objective, after successfully activating tumor-specific T cells [29], is for antigen-specific T cells to bind to tumor cells and induce cytotoxicity [30]. Nonetheless, this phenomenological assumption previously has been used and validated with experimental data in several modeling studies [13, 22, 31–33]. Therefore, given our limited information about how cancer progresses over time in different patients at different disease stages, we consider the dePillis-Radunskaya Law a reasonable approach to modeling the killing of tumor cells. Also, we assumed the logistic functional form to represent tumor growth.

Usually, the choice of a tumor growth function in general is determined by clinical data, which can vary depending on the type of tumor, location, and disease stage, among other factors [34]. Several studies [13, 22, 31–34] have suggested that the logistic and Gompertz functions are the most suitable for modeling tumor growth. Both functions have exhibited good predictions when calibrated with experimental data, usually from animal models of breast, lung, and melanoma cancers [13, 22, 31–34]. Our choice was based on the simplicity of the logistic function, as it only requires the estimation of two, rather three parameters in the Gompertz function, and the lack of longitudinal clinical data.

The model was calibrated through model fitting using individual patient data from a clinical trial study [26]. We fitted the model on parameters *c, c*_4_, *d*, and *λ* (see Table 1). These model parameters describe the killing and proliferation interactions between activated T cells and tumor cells, which were modeled based on phenomenological assumptions. We employed global optimization to find the best fit model parameter values to individual patient data. We selected adjusted *R*^2^ as our goodness-of-fir measure. The adjusted *R*^2^ ranged from 0.74 to 0.95 across the model fits for the six patients. Model fits to patient 1, 5, and 6 data resulted with a lower adjusted *R*^2^ (0.7) which may be due to fewer number of data points compared to the rest of the patients (6 vs. 7 data points). Patient 5 data had the lowest adjusted *R*^2^ and it may be partially attributable to a much earlier peak T cell response comparing to the rest of the patients.

Our model was used to show some possible applications, such as the longitudinal tumor size change forecast for individual patients undergoing melanoma treatment and the prediction of clinical outcome given initial tumor stage (size) when receiving the vaccine. Using available information on the individual patient’s tumor stage and clinical history, we predicted that the tumor size (diameter) 200 days after the first vaccination for all six patients. The predicted tumor size of Patients 1, 3, 4, and 5 who had no report of disease recurrence on day 200 fell under 1 mm in diameter, which is clinically declared a “thin” melanoma, and is considered a favorable outcome with a low risk of cancer spreading [35]. This model prediction is consistent with the clinical reports of those patients. For Patients 2 and 6, we compared model predicted tumor size with measurement from CT scans. Though model predicted tumor size for Patients 2 and 6 was slightly off from CT scans measurement, the model predicted increases of tumor sizes for these two patients at day 200 illustrating their recurrence.

Moreover, this kind of model can also be used to compare different clinical options. For example, we used our model to explore the impact of vaccine treatment initiation after resection surgery on tumor progression. Our results showed that the best-case scenario for treatment initiation is when patients start vaccination with a tumor of less than or equal to 6.4 mm in diameter (Fig 6A-B). In Fig 6B, we observed that 90% of the patients achieved a tumor diameter of less than 1 mm at the end of treatment (favorable lesion); whereas in Fig 6A, 100% of patients’ tumor size fell under 1 mm. Therefore, the ‘best’ clinical outcome of a personalized neoantigen peptide vaccine is when a patient starts treatment with a tumor diameter size of less than 6.4 mm (1 *×* 10^7^ cells).

Sensitivity analysis of our model using estimated parameters revealed the parameters which have the greatest impact on total activated T cell count during vaccine treatment, as well as final tumor size. The model variable for activated T cells (*A*_*TC*_) was found to be highly sensitive throughout the treatment period (days 25, 112, 147) for all patients, to parameter *c*_4_ (maximum CD4^+^ T cell production rate stimulated by the interactions with tumor cells). For patients 1, 2, and 6, the variable *A*_*TC*_ was shown to be sensitive to the parameter *c* (maximum CD8^+^ T cell recruitment rate stimulated by interactions with tumor cells). Proliferation (or recruitment) of immune cells to the tumor site has been shown to be a favorable prognostic sign [36]. Consistently, parameters in our model involved in the proliferation/recruitment of immune cells show great influence on the efficiency of the immunotherapy in eliciting an immune response. Also, for all patients, both variables, activated T cells (*A*_*TC*_) and tumor cells (*T*) were shown to be sensitive to the parameter *d* (maximal killing rate by activated CD8^+^ T cells. This result shows the importance of this parameter, as it may help determine how effective an immunotherapy can be in triggering the immune system to elicit tumor cytotoxicity.

The parametrization of our model was limited by the small number of patients from the clinical study used for model calibration, their limited follow-up times and lack of individual level longitudinal tumor cell count (or size) data. Potential future accumulation of patient data would provide the opportunity for better characterization of patient-specific model parameters that can be tumor-stage specific. Both individual patient longitudinal tumor size and T cell response measurement might greatly improve model calibration and validation for a better clinical outcome prediction. However, we observed significant differences in parameters *a*_1_ and *a*_2_ which determine the efficiency of the immune system to recognize tumor cells and elicit an immune response. For example, the parameter values of *a*_1_ and *a*_2_ obtained by fitting the ‘no recurrence’ patients’ data to our model were substantially smaller compared to the values of *a*_1_ and *a*_2_ obtained through the fitting of the ‘recurrence’ patients’ data. Another limitation of our current parametrization approach is that certain parameters found in the literature may have associated uncertainty that could affect model outcome, such as errors in measurements or natural variations. Though this variance in the parameters is not always reported in the literature, we briefly explored how model variables of interest are affected by potential variations of these parameters through global sensitivity analysis (see S1 Appendix; section B.8). It was found that both model variables of interest, activated T cells and tumor cells, are highly sensitive to parameters *α*_*p*_, Λ, and *δ*_*M*_ (internalization rate of peptides by DCs, maximum growth and death rate of mature DCs, respectively). In addition, the model variable for the activated T cell was found to be highly sensitive to *b*_4_, *σ*_4_, *µ*_4_, and *µ*_8_ (maximum growth of naïve CD4^+^ T cells, maximum activation rate of CD4+ T cells, and death rate of activated CD4^+^ and CD8^+^ T cells, respectively). The model variable for tumor cells, also was found to be highly sensitive to *b*_8_, *σ*_8_, and *ρ*_8_ (maximum growth of naïve CD8^+^ T cells, maximum activation rate CD8^+^ T cells, and proliferation rate for activated CD8^+^ T cells).

Other limitations of our model include the consideration of key mechanisms related to the efficacy of the vaccine. Although some recent studies [30, 37] showed that CD4^+^ T cells may act directly and indirectly in killing tumor cells, we did not incorporate this mechanism into our model. Our model incorporates the well-known mechanism of CD4^+^ T cells helping in the activation of CD8^+^ cytotoxic T cells in the presence of tumor cells. However, future versions of this model may incorporate the assumption of CD4^+^ T cell’s direct cytotoxicity against tumor cells if further evidence becomes available. We have not considered the tumor antigen escape phenomenon, and thus cannot model the potential outcomes if cancer cells stop expressing the neoantigens used to formulate the vaccines. The current version of our model also does not feature, concomitant therapies used against malignancies, as is usually the case in real-life scenarios. Additionally, we have not incorporated T cell sub-types (e.g., memory, regulatory) or antibody-mediated tumor eradication explicitly. We recognize that these are all important considerations for our future work.

Another important potential addition to our model would be the incorporation of a post-vaccine therapy, such as a checkpoint inhibitor, to enable continued tracking of the patient’s disease progression and determine possible safety and efficacy outcomes of individual or combined treatments. Last, our model is not readily applicable in studying efficacy of personalized neoantigen vaccines indicated in malignancies of the brain due to disease-specific constraints and the existence of the blood-brain barrier.

The current stage of the model serves as a proof of concept displaying its utility in clinical outcomes prediction, lays foundation for developing *in silico* clinical trials, and aids in the efficacy assessment of personalized vaccines. This model is flexible in the sense that its structure may be modified to capture other key mechanisms involved in the interactions of the human immune system and personalized cancer immunotherapy, and it allows for exploration and quantification of the impact of therapy initiation times on trial outcomes.

## Methods

### Mathematical model

We developed a mechanistic model to study the interactive dynamics of a personalized neoantigen peptide cancer vaccine, the human immune system, and tumor cells. The proposed model consists of a system of coupled non-linear ordinary differential equations (ODEs) representing important molecular and cellular populations which depict biological mechanisms that embody essential aspects of a neoantigen vaccine and target patient cohorts. The model is composed of a single immune system compartment encompassing the implicit connection of the innate and adaptive immunity. We consider two modeling levels, the cellular and subcellular (or molecular). At the cellular level, we have cell populations such as, antigen presenting cells (APCs), naïve and activated CD4^+^ and CD8^+^ T cells (helper and cytotoxic T cells, respectively), and tumor cells in the patient. One of the critical steps in vaccination is the efficient presentation of the tumor-specific antigen (TSA) to the T cells. Dendritic cells (DCs) are the most efficient APCs and we parametrized our APC model based on DCs. APCs capture, process and present the antigens to the T cells using cell surface proteins, i.e., MHC class I and II molecules. Engagement of the MHC-antigen complex with the T cell receptors (TCRs), initiates the differentiation of antigen-specific T cells into effector T cells [38]. We model these mechanisms and interactions at the cellular and subcellular levels.

Below, we describe in detail the assumptions considered for each equation with the end goal of constructing the full model. A schematic diagram of the mechanisms and interactions considered are shown in Fig 7 (cellular level) and Fig 8 (subcellular level). A list of model state variables with their respective definitions and units is presented in Table 2.

**Table 2.** Model equation variables with definition and units. Subscript *s* = *j* for HLA-I allele and *s* = *k* for HLA-II allele

|  | Variable | Description | Unit |
| --- | --- | --- | --- |
| Cellular | $p$ | Peptide concentration in a single vaccine | pmol |
| | $A_d$ | Adjuvant concentration in a single vaccine | mg/L |
| | $D_I$ | Immature dendritic cell population | cells |
| | $D_M$ | Mature dendritic cell population | cells |
| | $N_{CD4}$ | Naïve CD4<sup>+</sup> T cell population | cells |
| | $A_{CD4}$ | Activated CD4<sup>+</sup> T cell population | cells |
| | $N_{CD8}$ | Naïve CD8<sup>+</sup> T cell population | cells |
| | $A_{CD8}$ | Activated CD8<sup>+</sup> T cell population | cells |
| | $T$ | Tumor cell population | cells |
| Subcellular | $p^E$ | Amount of endosomal peptide fragments | pmol |
| | $M_s^E$ | Amount of endosomal MHC-I or MHC-II | pmol |
| | $pM_s^E$ | Amount of endosomal p-MHCI/II complex | pmol |
| | $pM_s$ | Amount of p-MHCI/II complex on DC membrane | pmol |
| | $M_s$ | Amount of free MHC-I/II on DC membrane | pmol |

**Fig 7.**
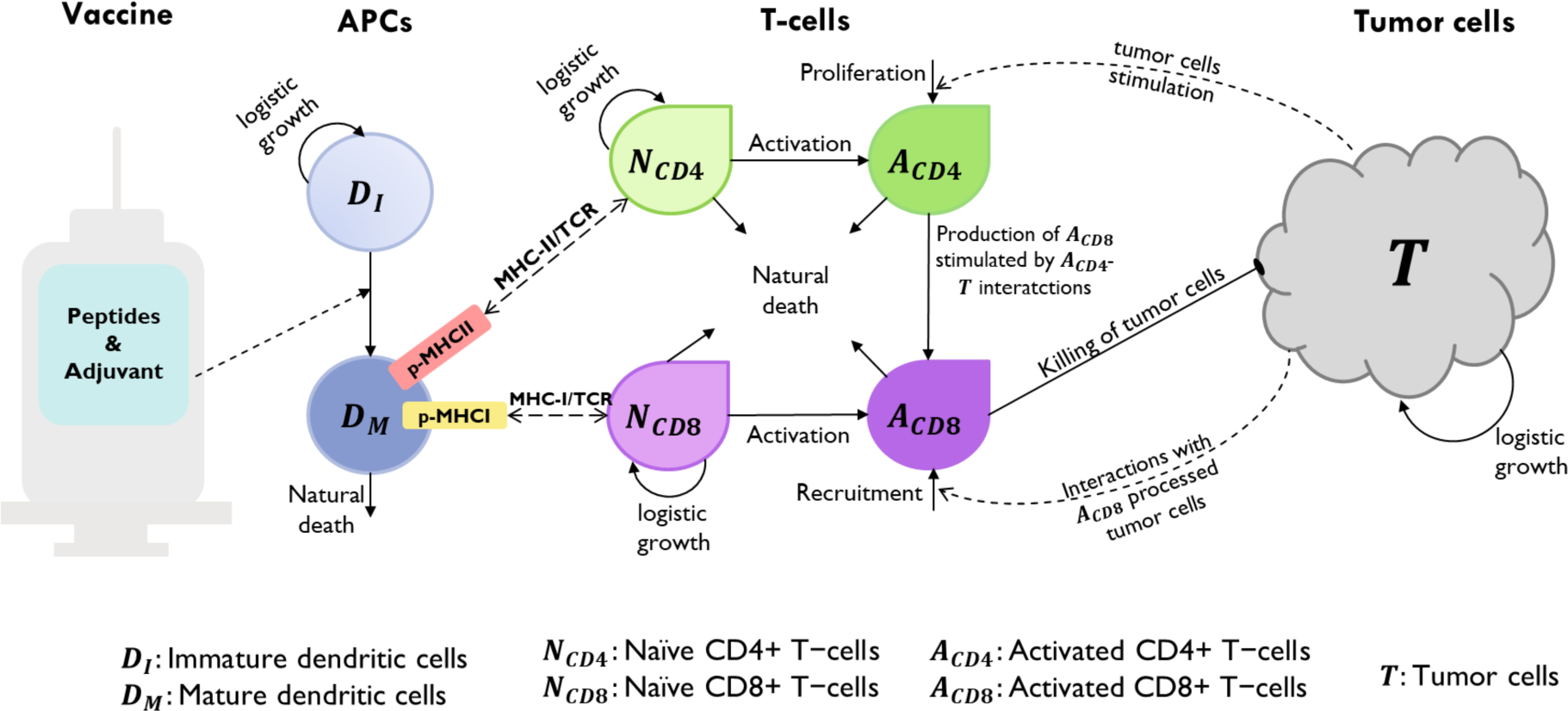
Schematic diagram of proposed mathematical model of the interactions between a human immune system and neoantigen cancer vaccines at the cellular level. Immature dendritic cells differentiate into mature dendritic cells due to the maturation signal provided by the adjuvant (dotted arrow). Upon antigen uptake, processing and presentation by the mature dendritic cells, naïve T cells recognize these (dashed lines with double-ended arrow-head) and differentiate into activated T cells that proliferate. Activated CD8^+^ T cells are now capable of killing tumor cells. Tumor lysis increase activated T cell counts through proliferation/recruitment (dotted curved arrows).

**Fig 8.**
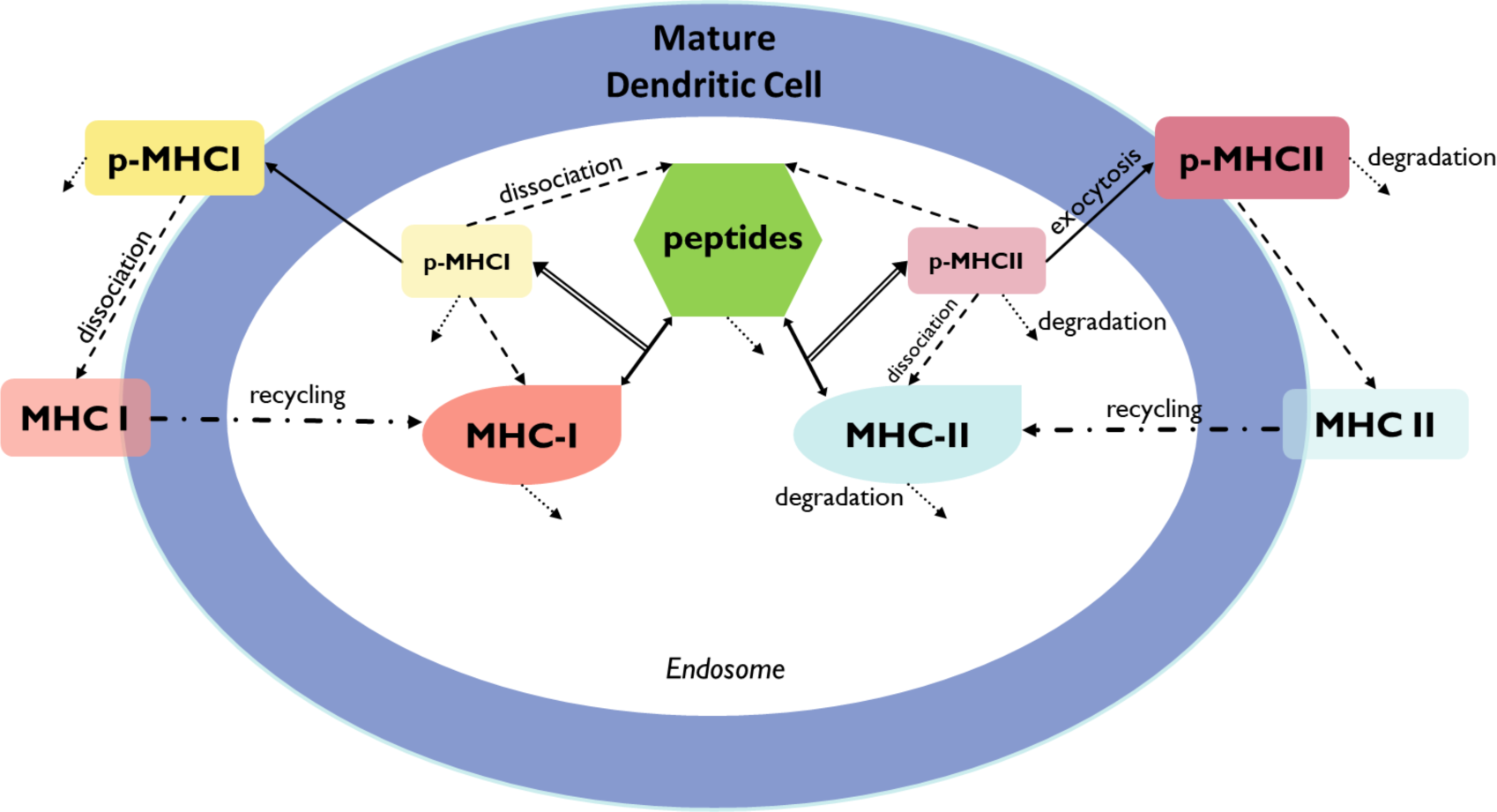
Schematic diagram of the mathematical model at the subcellular level. Mature DCs uptake peptides, which are then processed through the interaction with major histocompatibility complex (MHC) class I and II to produce peptide-MHC complexes (p-MHC). These p-MHCs are then presented in the DC membrane for further recognition by naïve T cells. Peptides, MHC and p-MHC complexes can suffer degradation. Peptide-MHC complexes have the potential to dissociate back into their constituents. MHC molecules on the DC membrane can be recycled back into the endosome. Double line arrows connecting to double head arrows represent the interaction of peptide with MHC-I or MHC-II molecules to create p-MHCI or p-MHCII complexes. Short dotted arrows represent degradation for peptides, MHC-I/II, and p-MHCI/II in the endosome and on the cell membrane. Dashed arrows represent dissociation of peptide-MHCs into peptide and MHCs molecules. Dash-dotted arrows represent the recycle of MHC complexes into the endosome after dissociation of p-MHCs on the cell membrane.

### The vaccine

The vaccine dosage in the model is composed of a certain concentration of an adjuvant and a load of peptides previously for a specific individual patient. Adjuvant is necessary to prime or ‘activate’ immature dendritic cells to respond to endogenous and exogenous antigens and elicit an immune response through the activation of T cells.

Let *p*(*t*) represent the peptide concentration in a vaccine and *A*_*d*_(*t*) the adjuvant concentration in a vaccine deposited in the subcutaneous (sc) connective tissue (injection site) at time *t*. We assume the vaccine concentration entering the tissue is instantaneously mixed and is homogeneously distributed for all *t*. Also, it is assumed that the vaccine is instantaneously absorbed by the body at time *t*, i.e., the vaccine’s concentration is updated before its content continues to be processed by dendritic cells. Since we consider that a vaccine dosage is administered at different scheduled time points, the Dirac delta function *δ* allows the description of bolus repeated administration of the same vaccine dose at times *τ*_*i*_, *i* = 1, …, *n*:

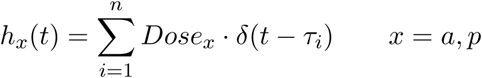

where *Dose*_*x*_, *x* = *p, a* is constant (specific to adjuvant or peptide concentration) and *τ*_*i*_ = 0, 3, 7, 14, 21, 83, 139 specifies the day when a vaccine is administered, with index *i* = 1, 2, …, 7 representing the number of vaccine doses administered throughout treatment. Note that the selection of these time points, *t*_*i*_, are based on the clinical study [26]. Thus, an equation representing the vaccine disposition process is:

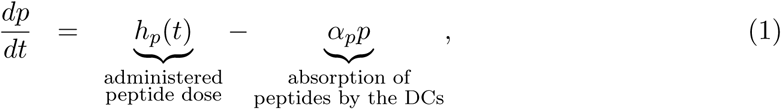

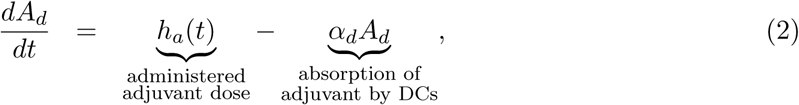

where *α*_*p*_ and *α*_*d*_ is the absorption rate of peptides and adjuvant by both immature and mature dendritic cells, respectively.

### Antigen-presenting cells: dendritic cells (DCs)

We assumed that immature dendritic cells, *D*_*I*_(*t*), located at the subcutaneous connective tissue grow logistically with a maximum rate Λ and carrying capacity *K*_*DC*_. Mature dendritic cells are assumed to die naturally with a rate *δ*_*M*_.

Immature dendritic cells, *D*_*I*_(*t*), can differentiate into mature dendritic cells, *D*_*M*_ (*t*), with a maximum differentiation rate of *r*_*D*_ in the absence of peptides and adjuvant. However, the maturation of DCs increases as a result of the contents of the vaccine. It is known that adjuvant increases the maturation of dendritic cells [39]. Thus, we consider an external maturation signal given by the effects of the adjuvant in the vaccine. In this case, the maturation signal is given by the release/absorption of adjuvant concentration, *A*_*d*_(*t*), in a vaccine, meaning, higher *A*_*d*_(*t*) concentration causes higher cell maturation, hence, larger mature dendritic cell, *D*_*M*_ (*t*), population size. When the concentration *A*_*d*_(*t*) is equal to *K*_*a*_, a half-maximum effect constant, the maturation rate becomes 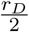. Given these biological assumptions, the equations describing the rate of change of immature dendritic cells, *D*_*I*_(*t*), and mature dendritic cells, *D*_*M*_ (*t*), are as follows:

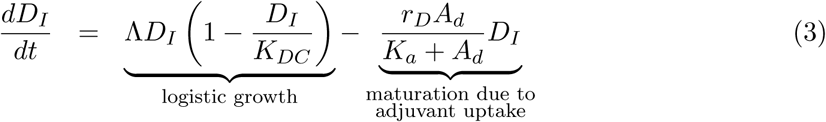

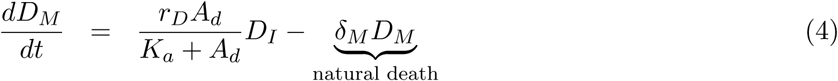

### Antigen processing and presentation by mature DCs

The following modeling approach follows a similar modeling framework of [40, 41]. The antigen processing and presenting by mature dendritic cells take place at the subcellular level (see Fig 8), in the endosomes of mature DCs. The vaccine peptides, *p*(*t*), are endocytosed into endosomes at a rate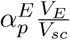, where 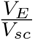 is a re-scaling factor of the volume of endosomes in a single DC, *V*_*E*_, and the volume of the injection site *V*_*sc*_. Therefore, peptides in DC’s endosomes are represented by a single molecular specie *p*^*E*^(*t*).

Inside the endosomes and other specialized vesicular compartments, peptides interact with MHC molecules of class I and/or class II. The strength of interaction of *p*^*E*^(*t*) and MHC molecule allele *s* is dictated by the effective disassociation constant 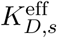 which gives the relationship between *k*_off,*s*_, the off-rate, and *k*_on,*s*_, the bio-molecular on-rate. The dissociation constant is defined as *K*_*D,s*_ = *k*_off,*s*_*/k*_on,*s*_.

Endosomal peptides, *p*^*E*^(*t*), bind to either MHC-I or MHC-II molecules in the endosomes, 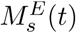 with allele *s* = *j* or *k*, at a rate *k*_on,*s*_, to form endosomal peptide-MHC-I (p-MHCI) or p-MHCII complexes, represented with 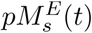. Note that binding occurs with available 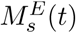 in the endosomes; thus, we consider a re-scaling factor of 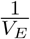. The p-MHCs may dissociate back into their respective components, peptide and MHC molecule, at a rate *k*_off,*s*_*/V*_*E*_. Also, the peptide fragments that cannot bind with an MHC molecule are further processed into its amino acids at rate *β*_*p*_.

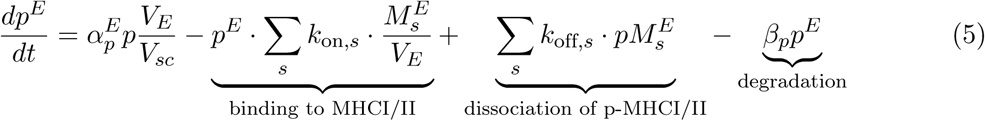

To calculate 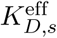, we follow a similar approach as in [40]. For each peptide-based vaccine sequence, we first obtain all *N* (15-30)-mer peptides and their individual MHC-I/II binding affinities *K*_*Dm*_, *m* = 1, …, *N*, from NetMHCpan 4.0 [42] and NetMHCIIpan 4.0 [43], respectively. Then, those (15-30)-mers are pooled into a single representative molecule that binds to MHC-I allele type *j* and an MHC-II allele type *k* with an ‘effective disassociation constant’ such that

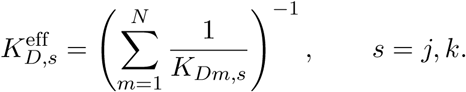

The molecular population dynamics of the endosomal free MHC-I/II molecules, 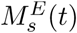, *s* = *j, k*, is given by Eq. (6), in which we assume: (1) homeostatic growth of endosomal free MHC-I/II molecules, with growth rate *β*_*M*_, and 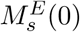 is the total number of MHC-I/II molecules at *t* = 0; (2) binding on and off between *p*^*E*^(*t*) and MHCI/II molecules in the endosomes, 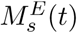; (3) recycling of free MHC-I/II from the cell membrane back into the DC’s endosomes with rate *k*_in_.

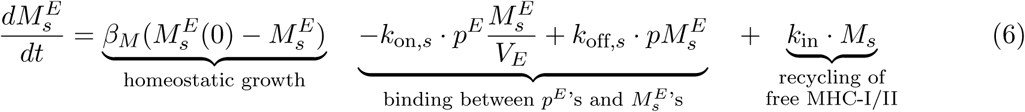

Equation (7) describes the molecular population dynamics of endosomal p-MHCI/II allele *j* or 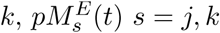, starting with the formation of p-MHCI/II complex. These can dissociate back into peptides and MHC components, degrade at a rate *β*_*pM*_, or get trafficked out at a rate *k*_ext_ to form membranal p-MHCs, *pM*_*s*_(*t*), which will be recognized by T cells.

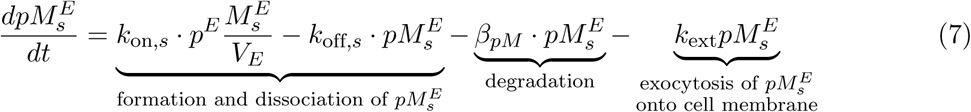

Moreover, the p-MHCs on the DC membrane can dissociate into free MHCs on the cell membranes, *M*_*s*_(*t*), at a rate *k*_off_ as in Eq. (8) and (9).

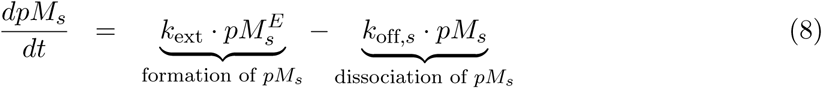

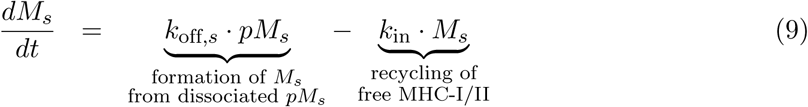

Let *pM*_*n*_, with *n* = I, II representing the MHC class I or II, denote the number of p-MHC molecules recognizable by the neoantigen-specific (CD4^+^ or CD8^+^) T cells, such that

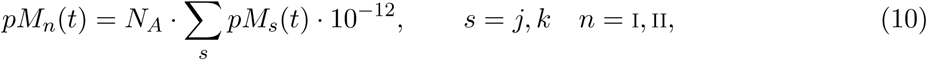

where *N*_*A*_ is Avogadro’s constant.

### Tumor cells

The total number of tumor cells at time *t, T* (*t*), is assumed to increase in a logistic fashion in the absence of immune intervention, where *r* is the maximum tumor growth rate and *K*_*T*_ is the tumor’s carrying capacity (maximum tumor cell burden). Activated CD8^+^ T cells kill tumor cells by a product of a maximum lysis rate *d*, and a rational expression that models tumor cell lysis as a function of the ratio of activated CD8^+^ T cells to tumor cells.

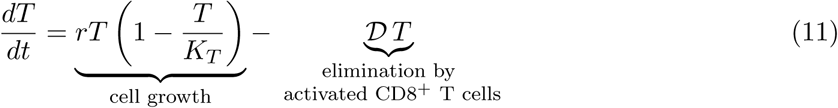

Where 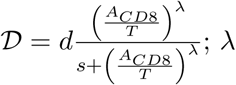 represents how the lysis rate depends on the effector/target ratio, and *s* affects the steepness of the curve, or how fast maximum killing rate is achieved. This expression is known as dePillis-Radunskaya Law (see [15, 31] for more details), which has been used extensively in the literature.

### Helper and cytotoxic T cells

Naïve CD4^+^ and CD8^+^ T cells are assumed to grow logistically (death regulated by carrying capacity), with maximum growth rate *b*_4_ and *b*_8_, and carrying capacity *K*_*TC*4_ and *K*_*TC*8_, respectively. Activated T cell population increases if there is a ‘successful’ activation process. Both naïve and activated T cells are assumed to die naturally with a rate *µ* (for both types of naïve T cells), *µ*_4_ (for activated CD4^+^ T cells), and *µ*_8_ (for activated CD8^+^ T cells).

The total number of naïve CD4^+^ and CD8^+^ T cells at time *t* is denoted with *N*_*CD*4_(*t*) and *N*_*CD*8_(*t*), respectively. Naïve T cells can recognize neoantigen fragments presented on mature DCs’ membrane (*pM*_*n*_, *n* = I, II) at any time *t*, represented with the product 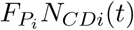, where 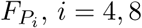, is the frequency of neoantigen-specific CD4^+^ (or CD8^+^) T cells. Naïve CD4^+^ and CD8^+^ T cells can be activated with a maximum activation rate of *σ*_4_ and *σ*_8_, respectively. The rate of change of naïve CD4^+^ (or CD8^+^) T cell population is modeled with the following equation:

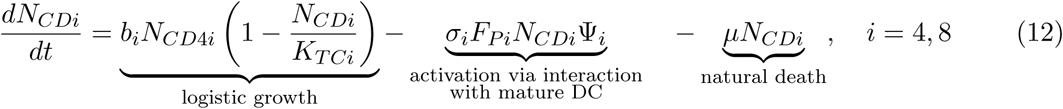

where, for example for *i* = 4, Ψ_4_ = Ψ_4_(*D*_*M*_, *N*_*CD*4_, *A*_*CD*4_), denotes the activation of neoantigen-specific precursor T cells dependent on the relative quantity of the mature dendritic cells to naïve and activated CD4^+^ T cells and the number of p-MHCII complexes,

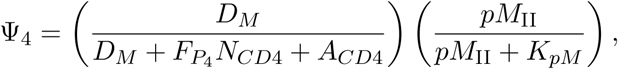

where *pM*_II_ represents the number of p-MHCII molecules recognizable by the neoantigen-specific CD4^+^ T cells as described in Eq. (10). The strength of mature DCs, *D*_*M*_ (*t*), to activate a neoantigen-specific CD4^+^ T cell is 50% when *pM*_II_(*t*) is equal to *K*_*pM*_ and increases as *pM*_II_(*t*) increases.

The total number of activated T cells is given by the equation:

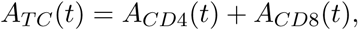

where *A*_*CD*4_(*t*) and *A*_*CD*8_(*t*) denote the population of activated CD4^+^ and CD8^+^ T cells, respectively. These cell populations increase with the activation of naïve CD4^+^ (or CD8^+^) T cells that had a ‘successful’ interaction with a mature DC. Activated T cells undergo proliferation or differentiation, depending on the proportion between *D*_*M*_ (*t*), cell types involved in the activation (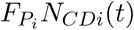, *D*_*M*_ (*t*) and *A*_*CDi*_(*t*)), and the number of T-epitope-MHC on *D*_*M*_ (*t*)’s, i.e., *pM*_I_(*t*) or *pM*_II_(*t*). The interaction of mature dendritic cells and activated CD4^+^ (or CD8^+^) T cells is described with the function:

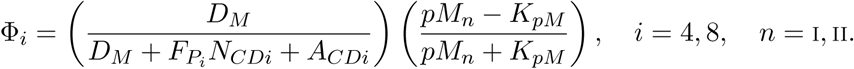

To understand this interaction better, for example, when the quantity *pM*_II_(*t*) at any time *t* is much larger than *K*_pM_, then the ratio 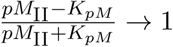, i.e., most cells proliferate. In a similar way, when *pM* _II_(*t*) is much smaller than *K_pM_*, then the ratio 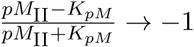, which implies most activated CD4^+^ T cells differentiate (i.e., cells are removed from this species compartment). The rate of change of activated CD4^+^ T cell population is modeled with the following equation:

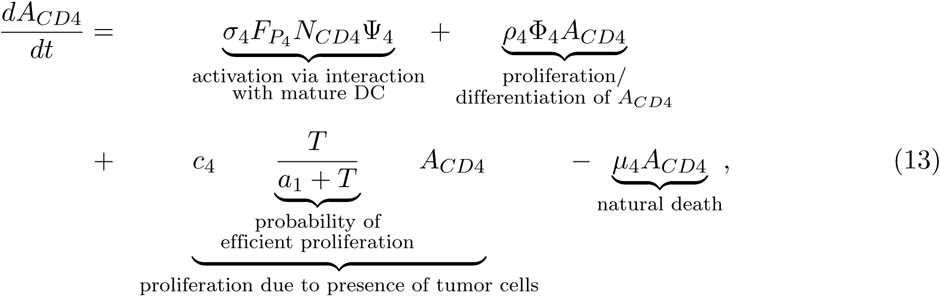

We assume an activating effect of CD4^+^ T cells due to existence of tumor cells represented by the third term in equation (13). This immune system’s response is described mathematically with a Michaelis-Menten interaction term, where the parameter *c*_4_ models the *antigenicity* of the tumor. Antigenicity can be thought of as a measure of how different the tumor is from ‘self’. This functional (or phenomenological) response to model this interaction has been used in other mathematical models such as [31, 44, 45].

The rate of change of activated CD8^+^ T cell population, *A*_*CD*8_(*t*), is modeled as follows:

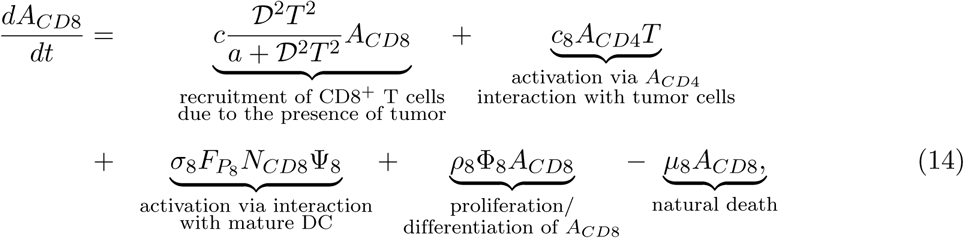

The first term of equation (14) represents the recruitment in CD8^+^ T cell population by interactions with T cell processed tumor cells through a Michaelis-Menten dynamic, as modeled before in [44, 46, 47]. Also, it is known that CD4^+^ T cells play a major role in providing help for anti-tumor cytotoxic T lymphocytes (CTLs) through direct mechanisms such as the secretion of interleukin (IL)-2, which directly activates CD8^+^ CTLs [30, 48]. We implicitly model this mechanism (second term) by assuming the activation of CD8^+^ T cells is given by Holling Type I functional response, *c*_8_*A*_*CD*4_*T*, where *c*_8_ is the rate at which CD8^+^ T cells are stimulated to be produced as result of activated CD4^+^ T cells interacting with tumor cells. The last three terms of equation (14) are similar to those described previously for the equation of *A*_*CD*4_(*t*).

### Model initial conditions and parameter values

Our mathematical model specifies 18 initial conditions such as immune and malignant cell counts, neoantigen peptide concentrations, and adjuvant concentrations in a vaccine (S1 Table). We categorized the initial conditions into two groups, ‘global’ and ‘patient-specific’. The global initial conditions (14 in total) are values that were found in the literature or estimated from available population level data of several clinical or experimental studies which include: immature and mature DC counts, naïve T cell counts, concentrations of endosomal peptides and MHC/p-MHC I and II molecules in DC’s endosome and on DC membrane. Patient-specific initial conditions (four in total) were estimated from patient-specific data from six patients in a personalized neoantigen anti-melanoma vaccine clinical trial (ClinicalTrials.gov: NCT01970358) [26]. These patient-specific initial conditions are the peptide and adjuvant concentrations in a vaccine, initial activated T cell counts, and initial tumor cell count. Detailed derivation of these estimates is provided in the Supplementary S1 Appendix.

Additionally, the model requires values for 45 parameters. A total of four model parameters were directly estimated by fitting the analytical solution of the total activated T cells, *A*_*TC*_(*t*) of six patients. These parameters are *c*: maximum CD8^+^ T cell recruitment rate; *c*_4_: maximum CD4^+^ T cell production rate; *d*: maximum lysis rate by activated T cells; *λ*: dependence of lysis rate on the effector/target ratio constant. We used the ‘NonlinearModelFit’ function in Mathematica Version 12.0 [49] with a constrained global optimization method (‘NMinimize’ with ‘SimulatedAnnealing’) for model parametrization. The goodness-of-fit was measured by the adjusted *R*^2^. Other parameters such as peptide and adjuvant concentration in the vaccine and patient-specific HLA alleles, were predetermined based on information provided in [26].

The detailed description of model parameters, parameter values/ranges found in the literature, parameters estimates from model fitting and parameter values used to run our simulations were organized by cell and molecule species on the Supplementary S1 Table and marked accordingly: parameters and initial conditions directly obtained from the literature were labeled with *♣*, those estimated from published data (clinical trial or experiment) were labeled with ♦, and patient-specific parameters fitted individually to six patients’ data from ClinicalTrials.gov: NCT01970358 were labeled with ⋆.

### Sensitivity analysis

We perform a global sensitivity analysis using Latin Hypercube Sampling (LHS) along with Partial Rank Correlation Coefficient (PRCC) to assess the sensitivity of two outputs of interest, activated T cells, *A*_*TC*_ and tumor cells, *T*, to the changes of the parameter values. In general, the magnitude of the PRCC indicates the impact of the uncertainty of estimating input parameter values for predicting the outcome variable, and the sign indicates the direction of correlation between each input parameter and each output variable [28, 50].

PRCC values were calculated between for each of the four patient-specific parameters (*c, c*_4_, *d, λ*), as well as two patient initial condition inputs *N*_*TC*0_ and *T*_0_ against two outputs of interest (*A*_*TC*_ and *T*) at specific time-points (days 25, 112, and 147). Through uncertainty quantification and sensitivity analysis, we determined which input parameters (see Table 1 for a list of estimated parameter values and Table 2 and the Supplementary S1 Table for definitions of the model variables and list of baseline parameters) are most influential on the outcome variables of interest. We used the mean of the best-fit parameter values of all six patients in Table 1 as the baseline to calculate LHS/PRCC values and the minimum and maximum of the best-fit parameter values to calculate the range of LHS/PRCC. Vaccine administration for all patients in the clinical trial [26] included a priming and a booster phase. We selected day 25 (*t* = 24 in our simulation), to define the ‘priming phase’, since the last priming vaccination in [26] occurred about 3 weeks in; day 112 (*t* = 111) as the ‘peak T cell response’ of activated T cells (as observed in [26]); and day 147 (*t* = 146) as the ‘end of treatment’ since the last booster vaccination occurred on week 20.

## Data Availability

NA

## Acknowledgments

This project was supported in part by an appointment to the Research Participation Program at OBE/CBER, U.S. Food and Drug Administration, administered by the Oak Ridge Institute for Science and Education through an interagency agreement between the U.S. Department of Energy and FDA. The authors received no financial support from any source, and there is no conflict of interest. The authors thank Joanne Berger, FDA Library, for manuscript editing assistance. We also extend our gratitude to Dr. Angela M. Jimenez Valencia (FDA/CVM) for the discussions that helped improve this manuscript.

## Author Contributions

**Conceptualization:** MRM, ONY, ZES, HY

**Data curation:** MRM

**Formal analysis:** MRM

**Funding acquisition:** HY

**Investigation:** MRM, ONY

**Methodology:** MRM, ONY

**Project administration:** MRM, ONY, HY

**Resources:** HY

**Supervision:** HY, ZES

**Visualization:** MRM

**Writing – Original draft:** MRM, ONY, ZES, UN, JRM, HY

**Writing – Review** & **Editing:** MRM, ONY, ZES, UN, JRM, HY

## Disclaimer

This article reflects the views of the authors and should not be construed to represent FDA’s views or policies.

## Supplementary information

**S1 Appendix. Mathematical details, parameter estimation, table of model parameter values and definitions, and additional results**.

**S1 Table. Parameter values and definition**. Mathematical model parameter definitions, value ranges, and values used in simulations.

**S1 Code. Mathematica Notebook**. The code to the replicate time series simulations of the presented model is available for review in https://github.com/marisabel19/CancerVaxModel2021.

## References

1. Hollingsworth RE, Jansen K. Turning the corner on therapeutic cancer vaccines. NPJ Vaccines. 2019;4:7. doi:10.1038/s41541-019-0103-y.

2. Peng M, Mo Y, Wang Y, Wu P, Zhang Y, Xiong F, et al. Neoantigen vaccine: an emerging tumor immunotherapy. Mol Cancer. 2019;18(1):128. doi:10.1186/s12943-019-1055-6.

3. Bitton RJ, Guthmann MD, Gabri MR, Carnero AJL, Alonso DF, Fainboim L, et al. Cancer vaccines: An update with special focus on ganglioside antigens (Review). Oncol Rep. 2002;9(2):267–276. doi:10.3892/or.9.2.267.

4. Thomas S, Prendergast GC. Cancer Vaccines: A Brief Overview. Methods Mol Biol. 2016;1403:755–61. doi:10.1007/978-1-4939-3387-743.

5. Aldous AR, Dong JZ. Personalized neoantigen vaccines: A new approach to cancer immunotherapy. Bioorg Med Chem. 2018;26(10):2842–2849. doi:10.1016/j.bmc.2017.10.021.

6. Aurisicchio L, Pallocca M, Ciliberto G, Palombo F. The perfect personalized cancer therapy: cancer vaccines against neoantigens. J Exp Clin Cancer Res. 2018;37(1):86. doi:10.1186/s13046-018-0751-1.

7. Hausser J, Szekely P, Bar N, Zimmer A, Sheftel H, Caldas C, et al. Tumor diversity and the trade-off between universal cancer tasks. Nat Commun. 2019;10(1):5423. doi:10.1038/s41467-019-13195-1.

8. Hu Z, Ott PA, Wu CJ. Towards personalized, tumour-specific, therapeutic vaccines for cancer. Nat Rev Immunol. 2018;18(3):168–182. doi:10.1038/nri.2017.131.

9. Jiang T, Shi T, Zhang H, Hu J, Song Y, Wei J, et al. Tumor neoantigens: from basic research to clinical applications. J Hematol Oncol. 2019;12(1):93. doi:10.1186/s13045-019-0787-5.

10. Guo Y, Lei K, Tang L. Neoantigen Vaccine Delivery for Personalized Anticancer Immunotherapy. Front Immunol. 2018;9:1499. doi:10.3389/fimmu.2018.01499.

11. Yarchoan M, Johnson r B A, Lutz ER, Laheru DA, Jaffee EM. Targeting neoantigens to augment antitumour immunity. Nat Rev Cancer. 2017;17(9):569. doi:10.1038/nrc.2017.74.

12. Adam JA, Bellomo N. A survey of models for tumor-immune system dynamics. Springer Science Business Media; 2012.

13. dePillis LG, Eladdadi A, Radunskaya AE. Modeling cancer-immune responses to therapy. J Pharmacokinet Pharmacodyn. 2014;41(5):461–78. doi:10.1007/s10928-014-9386-9.

14. Anderson ARA, Maini PK. Mathematical Oncology. Bull Math Biol. 2018;80(5):945–953. doi:10.1007/s11538-018-0423-5.

15. d’Onofrio A, Gandolfi A, D’Onofrio A. Mathematical Oncology 2013. Springer; 2014.

16. Barbolosi D, Ciccolini J, Lacarelle B, Barlesi F, Andre N. Computational oncology–mathematical modelling of drug regimens for precision medicine. Nat Rev Clin Oncol. 2016;13(4):242–54. doi:10.1038/nrclinonc.2015.204.

17. Victori P, Buffa FM. The many faces of mathematical modelling in oncology. Br J Radiol. 2019;92(1093):20180856. doi:10.1259/bjr.20180856.

18. Wilson S, Levy D. A mathematical model of the enhancement of tumor vaccine efficacy by immunotherapy. Bull Math Biol. 2012;74(7):1485–500. doi:10.1007/s11538-012-9722-4.

19. Agur Z, Halevi-Tobias K, Kogan Y, Shlagman O. Employing dynamical computational models for personalizing cancer immunotherapy. Expert Opin Biol Ther. 2016;16(11):1373–1385. doi:10.1080/14712598.2016.1223622.

20. Castiglione F, Piccoli B. Cancer immunotherapy, mathematical modeling and optimal control. J Theor Biol. 2007;247(4):723–32. doi:10.1016/j.jtbi.2007.04.003.

21. Peng H, Zhao W, Tan H, Ji Z, Li J, Li K, et al. Prediction of treatment efficacy for prostate cancer using a mathematical model. Sci Rep. 2016;6:21599. doi:10.1038/srep21599.

22. Radunskaya A, de Pillis L, Gallegos A. A Model of Dendritic Cell Therapy for Melanoma. Frontiers in Oncology. 2013;3(56). doi:10.3389/fonc.2013.00056.

23. Radunskaya KRWIT A E. Mathematical Modeling of Tumor Immune Interactions: A Closer Look at the Role of a PD-L1 Inhibitor in Cancer Immunotherapy. Spora: A Journal of Biomathematics. 2018;4(1):25–41. doi:http://doi.org/10.30707/SPORA4.1Radunskaya.

24. de Pillis LG, Gu W, Radunskaya AE. Mixed immunotherapy and chemotherapy of tumors: modeling, applications and biological interpretations. J Theor Biol. 2006;238(4):841–62. doi:10.1016/j.jtbi.2005.06.037.

25. Lai X, Friedman A. Combination therapy of cancer with cancer vaccine and immune checkpoint inhibitors: A mathematical model. PLoS One. 2017;12(5):e0178479. doi:10.1371/journal.pone.0178479.

26. Ott PA, Hu Z, Keskin DB, Shukla SA, Sun J, Bozym DJ, et al. An immunogenic personal neoantigen vaccine for patients with melanoma. Nature. 2017;547(7662):217–221. doi:10.1038/nature22991.

27. Stark PB. SticiGui, Onsophic, and Statistics W21. University of California, Berkeley. 2011;.

28. Blower SM, Dowlatabadi H. Sensitivity and uncertainty analysis of complex models of disease transmission: an HIV model, as an example. International Statistical Review/Revue Internationale de Statistique. 1994; p. 229–243.

29. Butterfield LH. Cancer vaccines. BMJ. 2015;350:h988. doi:10.1136/bmj.h988.

30. Tay RE, Richardson EK, Toh HC. Revisiting the role of CD4+ T cells in cancer immunotherapy—new insights into old paradigms. Cancer Gene Therapy. 2020; p. 1–13.

31. de Pillis LG, Radunskaya AE, Wiseman CL. A validated mathematical model of cell-mediated immune response to tumor growth. Cancer Res. 2005;65(17):7950–8. doi:10.1158/0008-5472.CAN-05-0564.

32. Vaghi C, Rodallec A, Fanciullino R, Ciccolini J, Mochel JP, Mastri M, et al. Population modeling of tumor growth curves and the reduced Gompertz model improve prediction of the age of experimental tumors. PLoS computational biology. 2020;16(2):e1007178.

33. Benzekry S, Lamont C, Beheshti A, Tracz A, Ebos JM, Hlatky L, et al. Classical mathematical models for description and prediction of experimental tumor growth. PLoS Comput Biol. 2014;10(8):e1003800.

34. Lopez AG, Seoane JM, Sanjuan MA. A validated mathematical model of tumor growth including tumor-host interaction, cell-mediated immune response and chemotherapy. Bull Math Biol. 2014;76(11):2884–906. doi:10.1007/s11538-014-0037-5.

35. Society AC. Melanoma Skin Cancer Stages; 2019. Available from: https://www.cancer.org/cancer/melanoma-skin-cancer/detection-diagnosis-staging/melanoma-skin-cancer-stages.html.

36. Gao JQ, Okada N, Mayumi T, Nakagawa S. Immune cell recruitment and cell-based system for cancer therapy. Pharmaceutical research. 2008;25(4):752–768.

37. Matsuzaki J, Tsuji T, Luescher IF, Shiku H, Mineno J, Okamoto S, et al. Direct tumor recognition by a human CD4+ T-cell subset potently mediates tumor growth inhibition and orchestrates anti-tumor immune responses. Scientific reports. 2015;5(1):1–14.

38. Palucka K, Banchereau J. Dendritic-Cell-Based Therapeutic Cancer Vaccines. Immunity. 2013;39(1):38–48. doi:10.1016/j.immuni.2013.07.004.

39. Walker J, Tough DF. Modification of TLR-induced activation of human dendritic cells by type I IFN: synergistic interaction with TLR4 but not TLR3 agonists. Eur J Immunol. 2006;36(7):1827–36. doi:10.1002/eji.200635854.

40. Yogurtcu ON, Sauna ZE, McGill JR, Tegenge MA, Yang H. TCPro: an In Silico Risk Assessment Tool for Biotherapeutic Protein Immunogenicity. The AAPS journal. 2019;21(5):96.

41. Chen X, Hickling T, Vicini P. A mechanistic, multiscale mathematical model of immunogenicity for therapeutic proteins: part 1—theoretical model. CPT: pharmacometrics & systems pharmacology. 2014;3(9):1–9.

42. Jurtz V, Paul S, Andreatta M, Marcatili P, Peters B, Nielsen M. NetMHCpan-4.0: improved peptide–MHC class I interaction predictions integrating eluted ligand and peptide binding affinity data. The Journal of Immunology. 2017;199(9):3360–3368.

43. Reynisson B, Barra C, Kaabinejadian S, Hildebrand WH, Peters B, Nielsen M. Improved prediction of MHC II antigen presentation through integration and motif deconvolution of mass spectrometry MHC eluted ligand data. Journal of Proteome Research. 2020;.

44. de Pillis LG, Gu W, Radunskaya AE. Mixed immunotherapy and chemotherapy of tumors: modeling, applications and biological interpretations. J Theor Biol. 2006;238(4):841–62. doi:10.1016/j.jtbi.2005.06.037.

45. de Pillis LG, Radunskaya A. - A mathematical model of immune response to tumor invasion. In: Bathe KJ, editor. Computational Fluid and Solid Mechanics 2003. Oxford: Elsevier Science Ltd; 2003. p. 1661 – 1668. Available from: http://www.sciencedirect.com/science/article/pii/B9780080440460504048.

46. de Pillis LG, Radunskaya A. - Modeling Tumor–Immune Dynamics. In: Eladdadi A MD Kim P, editor. Mathematical Models of Tumor-Immune System Dynamics. New York, NY: Springer Proceedings in Mathematics Statistics; 2014. p. 59–108.

47. Ghaffari A, Bahmaie B, Nazari M. A mixed radiotherapy and chemotherapy model for treatment of cancer with metastasis. Mathematical methods in the applied sciences. 2016;39(15):4603–4617.

48. Borst J, Ahrends T, Babal-a N, Melief CJ, Kastenmüller W. CD4+ T cell help in cancer immunology and immunotherapy. Nature Reviews Immunology. 2018;18(10):635–647.

49. Mathematica, Version 12.0;. Available from: https://www.wolfram.com/mathematica.

50. Marino S, Hogue IB, Ray CJ, Kirschner DE. A methodology for performing global uncertainty and sensitivity analysis in systems biology. J Theor Biol. 2008;254(1):178–96. doi:10.1016/j.jtbi.2008.04.011.

