## Supplementary material for "Mathematical Model of a Personalized Neoantigen Cancer Vaccine and the Human Immune System: Evaluation of Efficacy": S1 Appendix

### S1 Appendix. Mathematical details, parameter estimation, and additional results

#### A. Brief mathematical analysis

Briefly, we present routinely used mathematical analysis to show that the system is positively invariant, that is, the solutions can never be negative. Dynamical systems theory states that if we start in a positively invariant subset of the state space from which the dynamics of the ODE are under consideration, then we are guaranteed to stay within the set no matter how long ($t \to \infty$) we follow the dynamics of the ODE system.

**Theorem 1.** *Assume all model parameters in Table* **??** *are positive. The system (1)-(15) is positive invariant in* $\mathbb{R}_+^{14}$.

*Proof.* We want to show that, if initial conditions of the state variables in our model are greater than 0, meaning, $p(0) > 0$, $A_d(0) > 0, D_I(0) > 0$, $D_M(0) > 0$, $p^E(0) > 0$, $M^E(0) > 0$, $pM^E(0) > 0$, $pM(0) > 0$, $M(0) > 0$, $N_{CD4}(0) > 0$, $N_{CD8}(0) > 0$, $A_{CD4}(0) > 0$, $A_{CD8}(0) > 0$, and $T(0) > 0$, then for all $t > 0$, solutions to each of the state variables stay in the positive quadrant, meaning, $p(t) > 0$, $A_d(t) > 0, D_I(t) > 0$, $D_M(t) > 0$, $p^E(t) > 0$, $M^E(t) > 0$, $pM^E(t) > 0$, $pM(t) > 0$, $M(t) > 0$, $N_{CD4}(t) > 0$, $N_{CD8}(t) > 0$, $A_{CD4}(t) > 0$, $A_{CD8}(t) > 0$, and $T(t) > 0$.

Thus, for any $p \geq 0$, $A_d \geq 0, D_I \geq 0$, $D_M \geq 0$, $p^E \geq 0$, $M^E \geq 0$, $pM^E \geq 0$, $pM \geq 0$, $M \geq 0$, $N_{CD4} \geq 0$, $N_{CD8} \geq 0$, $A_{CD4} \geq 0$, $A_{CD8} \geq 0$, and $T \geq 0$, whe have that

$$p'\Big|_{p=0} = Dose_p \geq 0, \qquad A_d'\Big|_{A_d=0} = Dose_a \geq 0, \qquad D_I'\Big|_{D_I=0} = 0,$$

$$D_M'\Big|_{D_M=0} = \frac{r_D A_d}{K_a + A_d} D_I \geq 0, \qquad N_{CD4}'\Big|_{N_{CD4}=0} = 0, \qquad N_{CD8}'\Big|_{N_{CD8}=0} = 0, \qquad T'\Big|_{T=0} = 0,$$

$$A_{CD4}'\Big|_{A_{CD4}=0} = \sigma_{Nh} F_{P1} N_{CD4} \Phi_1 \geq 0, \qquad A_{CD8}'\Big|_{A_{CD8}=0} = \sigma_{Nc} F_{P2} N_{CD8} \Phi_3 \geq 0,$$

$$p^{E'}\Big|_{p^E=0} = \alpha_p p \frac{V_E}{V_{sc}} \geq 0, \qquad M^{E'}\Big|_{M^E=0} = k_{\text{in}} M \geq 0, \qquad pM^{E'}\Big|_{pM^E=0} = k_{\text{on}} \cdot p^E \frac{M^E}{V_E} \geq 0$$

$$pM'\Big|_{pM=0} = k_{\text{ext}} \cdot pM^E \geq 0, \qquad M'\Big|_{M=0} = k_{\text{off}} \cdot pM \geq 0.$$

This implies that due to the continuity of the system, it is impossible for any of these state variable solutions to drop below 0. □

### B. Parameter Estimation

#### B.1. Estimating peptide concentration in a vaccine from amino acid sequences

We extracted the amino acid sequence of the peptides used for each patient's personalized vaccine used in the melanoma study [1] from their supplementary material. First, using all amino acid sequences of peptides in each vaccine, we calculated the protein molecular weight (KDa) by using the online tool *The Sequence Manipulation Suite: Protein Molecular Weight* [2]. Then, in [1], it was stated that each peptide had a weight of 0.3 mg. We used the online tool [3] to covert the weight concentration to pmol. In Table A1, we lay out each of the components necessary to calculate the peptide concentration, $Dose_p$ in pmol for input in our model.

**Table A1. Peptide dose conversion from *mg* to *pmol* by patient with melanoma in [1].**

| Patient | No. of peptides | Weight (mg) | Protein molecular weight (kDa) | Amount (pmol) |
|---|---|---|---|---|
| 1 | 13 | 3.9 | 32.68 | 119,340 |
| 2 | 17 | 5.1 | 42.49 | 120,030 |
| 3 | 14 | 4.2 | 38.33 | 109,570 |
| 4 | 14 | 4.2 | 36.03 | 116,570 |
| 5 | 20 | 6 | 54.12 | 110,860 |
| 6 | 20 | 6 | 53.66 | 111,820 |

#### B.2. Dendritic cells

**Carrying capacity, $K_{DC}$.** To estimate the carrying capacity of immature dendritic cells in the total volume of the subcutaneous tissue injection site, we used the total number of dendritic cells in Figure 4A of [4]. Since this article shows the total number of DCs by gender, we use the total number of DCs in females (approx. 27 number of cells/$\mu$L) as the lower bound of our range and we use the total number of DCs in males (approx. 35 number of cells/$\mu$L) as the upper bound. Estimating the number of DCs cells per volume we obtain:

$$27 \text{ to } 35 \quad \frac{\text{cells}}{\mu\text{L}} \cdot \frac{\mu\text{L}}{1 \times 10^{-6}\ \text{L}} = 27 \text{ to } 35 \times 10^{6}\,\frac{\text{cells}}{\text{L}}$$

Now, to estimate the number of cells at the injection site, we multiply by the total volume of the subcutaneous tissue ($V_{sc}$):

$$27 \text{ to } 35 \times 10^{6}\frac{\text{cells}}{\text{L}} \times 0.7676\ \text{L} = 2.07252 \text{ to } 2.6866 \times 10^{7} \text{ cells.}$$

Thus, we choose the median $2.38 \times 10^{7}$ as the carrying capacity of immature dendritic cells.

**Immature DCs maturation rate function: Half-maximum adjuvant effect constant, $K_a$, and maximum differentiation rate, $r_D$.** To estimate parameters $r_D$ and $K_a$, we fitted the analytical solution (see Eq. (1) below) of the second term in the equation of our model, $D_I'(t)$, representing the maturation rate of immature DCs to data from Figure 6A in [5] using Mathematica 12.0 [6] by looking for the best fit. The data from Figure 6A in [5] was extracted using WebPlotDigitizer [7]. Figure 6A in [5], shows how DCs mature as a function of poly(I:C) adjuvant. In this figure, authors use CD83 since it is known that these are activation markers for antigen presenting cells [8]. We re-scaled $\%\text{CD83}^+$ cells to proportion of matured DCs in the y-axis and microgram per milliliter to milligram per liter in the $x$-axis of Figure 6A resulting in Figure 1 (see below).

Using separation of variables method, we find the analytical solution required to estimate $r_D$ and $K_a$,

$$\frac{dD_I(t)}{dt} = -\frac{r_D A_d}{K_a + A_d} D_I(t)$$
$$\frac{1}{D_I(t)} dD_I(t) = -\frac{r_D A_d}{K_a + A_d}\, dt$$

and integrate such that for all $t \in [0, T)$,

$$\int_{D_I(0)}^{D_I(T)} \left(\frac{1}{D_I(t)}\right) dD_I(t) = \int_0^T -\frac{r_D A_d}{K_a + A_d}\, dt$$
$$\ln(D_I(T)) - \ln(D_I(0)) = -\frac{r_D A_d}{K_a + A_d} T$$
$$D_I(T) = D_I(0) \exp\left(-\frac{r_D A_d}{K_a + A_d} T\right)$$

Now, assuming $D_M(0) = 0$ and $D_M(T) = D_I(0) - D_I(T)$ such that $\frac{D_M(T)}{D_I(0)} + \frac{D_I(T)}{D_I(0)} = 1$ (i.e., proportion of immature and mature DCs at time $T$ equals to 1), then

$$\frac{D_M(T)}{D_I(0)} = 1 - \exp\left(-\frac{r_D A_d}{K_a + A_d} T\right)$$

According to the experiments done in [5], the cells were cultured for 24 hours before taking measurements of maturation. Hence, we set $T = 1$ (1 day) and the function to be fitted is:

$$f(x) = 1 - \exp\left(-\frac{r_D x}{K_a + x}\right) \tag{1}$$

Using the built-in function 'NonlinearModelFit' in Mathematica [6], we obtain the following parameter estimates from equation 1:

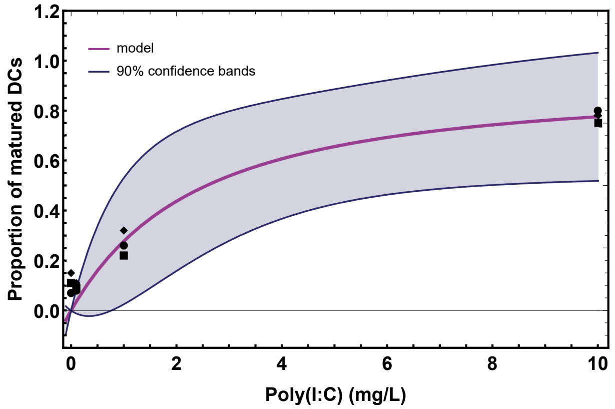


**Figure 1. Estimation of parameters $r_D$ and $K_a$.** Purple line represents the function: $f(x) = 1 - \exp\left(-\frac{r_D x}{K_a + x}\right)$, with point estimates $r_D = 2.48$ (SE: 1.43; 90%CI[0, 6.66]*) and $K_a = 6.64$ (SE: 6.37; 90%CI:[0, 25.26]*), fitted to data extracted from Figure 6A in [5]. 90% confidence bands for mean predictions are in blue. [90% CI]* denotes modified CI by replacing negative lower endpoint with zero as described in [9].

#### B.3. Estimation of parameters corresponding to the model equations that represent naïve T-cell populations

**Initial counts of naïve T-cells.** In Ott et al. [1], eligible patients needed to have a lymphocyte count of at least 800 cells per microliter. We use these number as a baseline to estimate naïve T-cell counts at $t = 0$ in our simulations. The average adult weighting 58 to 80 kilograms has about 4.5-5.7 L of blood. If we assume that patients have an average of 5 L of blood, then each patient has at least $4 \times 10^9$ lymphocytes. On average, using standard methods such as Ficoll-Paque density gradient centrifugation for PBMC yields 0.5-$2\times10^9$ cells/L of blood [10]. This will give us a range of $(2.5, 10) \times 10^9$ cells, with a median of $6.25 \times 10^9$ cells.

In Bittersohl et al. [10], authors provide percentages of different cells types which compose the PBMCs in a healthy adult human as shown in Table 9.1 of [10] (see Table A2 below). From this table, we obtain that 70% of PBMCs are lymphocytes for which 60 out of 70 percent (about 86% of total lymphocytes) are T-cells and 10 out of the 70 percent (about 14% of total lymphocytes) are B-cells. With these percentages, we obtain a T-cell range of $(2.15, 8.6) \times 10^9$ with a median of $5.38 \times 10^9$ T-cells. Since this median is larger than the baseline calculated above, we assign $5.38 \times 10^9$ cells as the initial naïve T-cell count in our simulations.

**T-cell Carrying capacity.** In order to have a one-to-one comparison of T-cell count from extracted data and simulation, we first estimate the average number of PBMCs in a human. Following [11], we assumed that a human has $1.22 \times 10^{12}$ lymphocytes. Using Table A2, we estimate there are $1.43 \times 10^{12}$ PBMCs. Thus, we conclude that T-cells $= 8.57 \times 10^{11}$ (60% of PBMCs).

**Table A2.** Table 9.1 from [10]

| **Cell Type** | **%** |
|---|---|
| T-cells ($CD3^+$) | 60 |
| Helper T-cells ($CD3^+$, $CD4^+$) | 70 of T-cells |
| Cytotoxic T-cells ($CD3^+$, $CD8^+$) | 30 of T-cells |
| B cells ($CD22^+$) | 10 |
| Monocytes/macrophages ($CD14^+$) | 15 |
| Natural killer (NK) cells ($CD56^+$)/$CD16^+$ | 15 |

#### B.4. IFN-$\gamma$ ELISPOT of PBMCs data extraction

In Ott el al. Figure 2b [1], the authors provided longitudinal plots for *ex-vivo* assay responses to peptide pools for each patient in the trial in units of per million PBMC. These data points were obtained by using an IFN-$\gamma$ ELISPOT assay. This method allows the quantification of the number of $CD4^+$ or $CD8^+$ T-cells which secrete IFN-$\gamma$ in response to a stimulation with a specific antigen or peptide [12, 13]. Moreover, each spot within an ELISPOT well identifies a single cell that has released a measurable amount of cytokine, i.e., the number of spots is a direct measure of the frequency of cytokine-producing T-cells in the PBMC population [12]. Thus, it is a reasonable assumption that the number of spots to number of cells is a one-to-one conversion.

To extract data from plots and images, we used WebPlotDigitizer [7] and plotted the extracted data (see Figure 2a) by patient. Each patient's frame contains the data of the four pools (Pool A - Pool D) which make up one vaccine dose and a baseline data (mock). Figure 2a is plotted in the form of cell counts by multiplying each extracted data point by $1.43 \times 10^6$ (see section: T-cell Carrying capacity), since each data point is in spot-forming cells per $10^6$ PBMCs. Then, to plot the total cell count of *ex-vivo* assay responses to peptides vaccine for each patient, we summed the data points of Pool A through D each subtracted by the Mock data at each time point and arrive at the numbers shown in Figure 2b. Due to the stochastic nature of *ex-vivo* assays, one caveat to subtracting the Mock data is that it may lead to negative cell counts on the treatment initiation day for which we assigned a small ($10^{-6}$) number of activated cells. Additionally, we assumed there is a +/- 15% variation to the ELISPOT counts presented by Ott et al., as it has been shown in [14] that the relative experimental error for 0-200 number of spots counted per well is between 0.1-0.2.

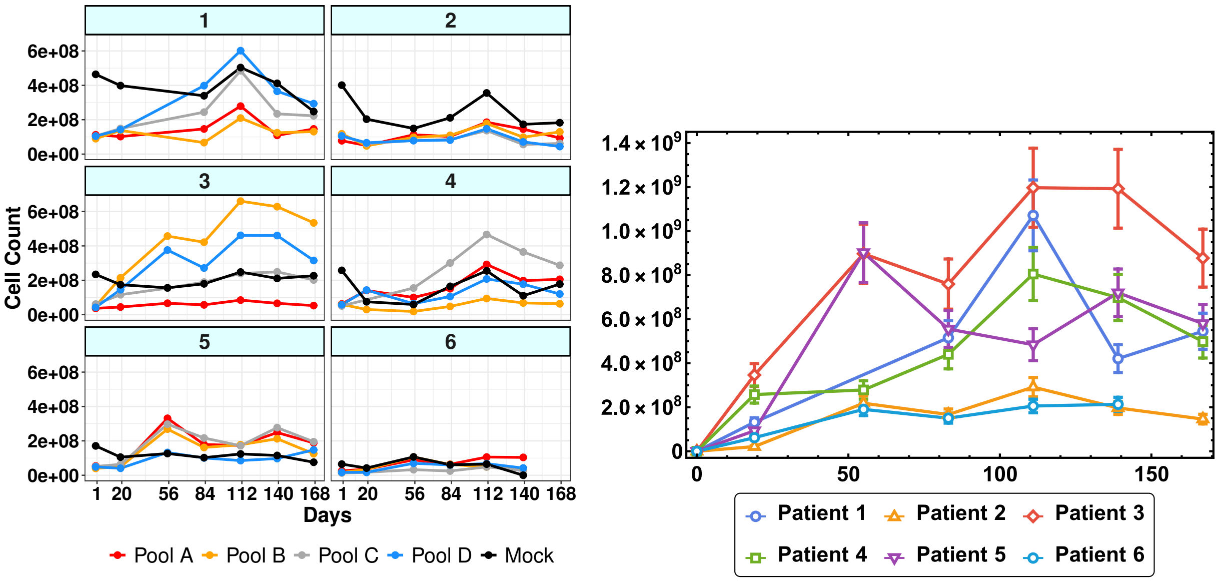


**(a)** Cell count of vaccine pools by patient

**(b)** Total cell count of vaccine by patient

**Figure 2.** Data transformation from *ex-vivo* IFN-$\gamma$ ELISPOT responses for PBMCs in [1] to cell count. (a) Cell count of T-cell response of each pool in a vaccine by patient. (b) Total cell count T-cell response of each patient's vaccine with melanoma.

**$CD4^+$ and $CD8^+$ T-cell initial counts.** The first data point at day 0 for each patient was used to assign to each patient's initial $CD4^+$ and $CD8^+$ counts. Given that these data only tell us the T-cell count without making distinction between cell subtypes, based on Table A2, we assume that T-cell counts on day 0 are distributed as 70% $CD4^+$ T-cells and 30% $CD8^+$ T-cells.

#### B.5. Parameters corresponding to the equation describing tumor cell population

**Estimating maximum cancer growth rate, $r$.** In Liu et al. [15], authors studied the rate of growth in melanomas, finding that one third of the melanomas grew at least 0.5 mm per month. The median monthly growth rate for nodular melanomas was 0.49 mm in diameter, superficial spreading melanomas at 0.12 mm in diameter, and lentigo melanomas at 0.13 mm/month [15]. The patients sample for our simulations in [1] were diagnosed at different stages of melanoma cancer (see Table A4). Four of the patients who did not have recurrence after vaccine initiation had nodular melanomas (i.e., the tumor has spread to nearby lymph nodes [16]). However, all patients before initiating vaccine treatment, underwent surgery, thus, we assumed the growth rate of any residual after surgery is 0.15 mm/month in diameter. That is, we are assuming the best case scenario for these patients, regardless their initial tumor stage diagnosis.

To determine the number of cancer cells of a tumor in a spherical shape, we considered that each cell is about 20 $\mu$m in diameter and a 1-mm cancer has about 100 thousand cells [17]. Therefore, to directly calculate the cell count in a tumor of diameter $d$, we used the formula of the volume of a sphere modified to incorporate a 25.95% of void volume [18], and multiply by

$1 \times 10^5$ cells:

$$\text{Total cell count in a spherical tumor: } T_c(d) = \frac{4\pi}{3}\left(\frac{d}{2}\right)^3 (0.7405) \cdot \frac{10^6\text{cells}}{\text{mm}^3}, \tag{2}$$

where $d$ is the diameter in millimeters (mm).

The tumor growth rate is then calculated as follows:

1. *Diameter per day:*
$$\frac{0.15 \text{ mm}}{30 \text{ days}} = \frac{0.005\text{mm}}{\text{day}}$$

2. *Tumor volume (void volume removed) per day:*
$$\frac{4\pi}{3}\left(\frac{0.004 \text{ mm}}{2}\right)^3 (0.7405) = 4.84 \times 10^{-8} \text{ mm}^3/\text{day}$$

3. *Total tumor cell count per day:*
$$4.84 \times 10^{-8} \text{ mm}^3/\text{day} \cdot 10^5 = 0.004 \text{ cells/day}$$

**Estimating initial tumor cell count, $T(0)$, by patient.** To estimate the initial tumor cell count on day 0 ($t = 0$), we used the information provided for each of the patients in the 'Extended Data Table 1: Characteristics of patients' along with the clinical event timeline from surgery until time of data cut off in 'Figure 1', both in [1]. Then, following the guidelines from the American Cancer Society [16] and Melanoma Research Alliance [19] for melanoma skin cancer staging and using nodal staging and metastasis information on [20] and [21], respectively, we made the following assumptions which are then summarized in Table A3:

(a) Given the tumor diameter ranges in [19] for each of the tumor thickness **(T)** in column 2 of Table A3, we make a definitive assumption for the tumor diameter.

(b) According to [20], lymph nodes are considered malignant if they measure more than 1 cm in the short axis diameter. Though, the size threshold varies with anatomic site and underlying tumour type. For example, in rectal cancer lymph nodes are considered pathological when they measure more than 5 mm. Hence we assigned (column 5 of Table A3) the number of tumor-involved nodes according to the code **(N)**.

(c) If a patient has metastasis **(M)**, then in [21] it was stated that lung metastasis ranges between 3-7 cm. Hence, we assumed that patients with metastasis will have three 5-cm in diameter lung metastasis (column 7).

As an example, we will walk you through the calculation to obtain the tumor cell count of Patient 2 at the beginning of cancer treatment (column 6 in Table A4):

| Tumor Stage | Tumor thickness at primary site | Assumption: size before surgery | Lymph nodes involvement | Assumption: no. of 5 mm tumor-involve nodes | Metastasis | Size: 3-7 cm |
|---|---|---|---|---|---|---|
| IIIC, IV | T2 > 1-2 mm | 2 mm | N0 (no regional metastasis) | 0 | M0 | 0 |
| | T3 > 2-4 mm | 4 mm | N2 (2-3) | 3 | M1b | 5cm (lung) |
| | T4 > 4 mm | 5 mm | N3 (4 or more) | 5 | | |

**Table A3. Definition of tumor diameter according to the American Joint Committee on Cancer (AJCC) TNM system [20].**

- Patient 2 in [1] was diagnosed with T4N0M1b. Using Table A3, we conclude that T4 corresponds to a tumor diameter of 5 mm; N0 means there are no tumor-involved nodes; M1b corresponds to one 50-mm lung metastasis. Then using equation (2), we compute the tumor cell count for a tumor diameter of 5 mm, adding three times the tumor cell count for a tumor of 50 mm in diameter,

$$T_c(5) + 3T_c(50).$$

Then, the tumor cell count after surgery is considered to be 5% of the tumor size before surgery. Thus, we multiply column 3 by 0.05. Lastly, we use the equation for tumor cells (Eq. (15) in the main text) without the second term which represents the intervention of the immune system. This means, we assume the tumor cells continue to grow without any mediation from the immune system.

$$\frac{dT}{d\tau} = 0.004\left(1 - \frac{T(\tau)}{1.45 \times 10^{10}}\right)T(\tau) \tag{3}$$

In the equation above, we use 0.004 as the maximum growth rate estimated previously and $1.45 \times 10^{10}$ as the tumor carrying capacity since this is the largest tumor cell count before surgery among patients (see column 2 in Table A4).

| Patient | Stage by tumor thickness | Tumor cell count BEFORE surgery | Tumor cell count AFTER surgery (5%) | Time lag (days) | Tumor cell count at the beginning of treatment |
|---|---|---|---|---|---|
| 1 | T3 (N3M0) | $1.702 \times 10^7$ | 851 056 | 120 | $1.375 \times 10^6$ |
| 2 | T4 (N0M1b) | $1.454 \times 10^{10}$ | $7.272 \times 10^8$ | 120 | $1.139 \times 10^9$ |
| 3 | T3 (N2M0) | $1.217 \times 10^7$ | 608 728 | 165 | $1.177 \times 10^6$ |
| 4 | T4 (N2M0) | $1.453 \times 10^7$ | 726 984 | 180 | $1.493 \times 10^6$ |
| 5 | T2 (N2M0) | $1.0 \times 10^7$ | 500 165 | 135 | 858 265 |
| 6 | T2 (N0M1b) | $1.454 \times 10^{10}$ | $7.27 \times 10^8$ | 105 | $1.078 \times 10^9$ |

**Table A4. Estimation of initial tumor cell count by patient at the beginning of vaccine treatment.** Third column is estimated using Table A. Fourth column is estimated using equation (2). Fifth column is the time lag between surgery and initiation of cancer vaccine treatment. Sixth column is estimated using equation (3).

#### B.6. On-rate for T-epitope-MHCII binding, $k_{\mathbf{on},2}$

In [22], it is described that the maximum on-rate constant for binding is approximately $10^5$ to $10^6$ $\mathrm{M}^{-1}\mathrm{s}^{-1}$, i.e.,

$$\frac{(10^5 \text{ to } 10^6)}{\mathrm{M}\cdot\ \mathrm{s}} \cdot \frac{10^{-12}\mathrm{M}}{\mathrm{pM}} \cdot \frac{86400\mathrm{s}}{\mathrm{day}} = (8.64 \times 10^{-3} \text{ to } 8.64 \times 10^{-2})\ \mathrm{pM}^{-1} \cdot \mathrm{s}^{-1}.$$

#### B.7. Detailed methodology for global sensitivity analysis

We chose $N = 1000$ as the number of LHS/PRC simulations. Following the approach and algorithm described in [23], we constructed an LHS matrix of size $N$ by $K$ ($N$: no. of rows; $K$: no. of columns). Hence, each row corresponds to one simulation and contains a randomly selected and equally distributed value for each of the uncertain parameters of interest. Each of these rows in the LHS matrix is then used to calculate each of the outcome variables of interest producing $N$ observations that are then used to assess the sensitivity of the outcome variables to the uncertainty in the input parameters. Next, we calculate the PRCC value for each outcome variable at specific time-points of interest ($t = 24$, 111, 146) following the approach of [23]. Sensitivity analysis simulations were performed in Mathematica, Version 12.0 [6].

#### B.8. Additional uncertainty and sensitivity analysis for model outputs

We briefly present the results of the global sensitivity analysis performed for two model variables of interest, activated T-cells and tumor cells, using parameters found in the literature which may have associated uncertainty that could affect model outcome, such as error in measurements or natural variations. It was found that both model variables of interest, activated T-cells and tumor cells, are highly sensitive to parameters $\alpha_p$, $\Lambda$ and $\delta_M$ (internalization rate of peptides by DCs, maximum growth and death rate of mature DCs, respectively). The sensitivity produced by these parameters are shown to decrease throughout time. Individually, in addition to $\alpha_p$, $\Lambda$ and $\delta_M$, model variable for activated T-cell was found to be highly sensitive to $b_4$, $\sigma_4$, $\mu_4$ and $\mu_8$ (maximum growth of naïve $CD4^+$ T-cells, maximum activation rate of $CD4^+$ T-cells, and death rate of activated $CD4^+$ and $CD8^+$ T-cells, respectively). The model variable for tumor cells, was found to be highly sensitive $b_8$, $\sigma_8$ and $\rho_8$ (maximum growth of naïve $CD8^+$ T-cells, maximum activation rate $CD8^+$ T-cells, and proliferation rate for activated $CD8^+$ T-cells).

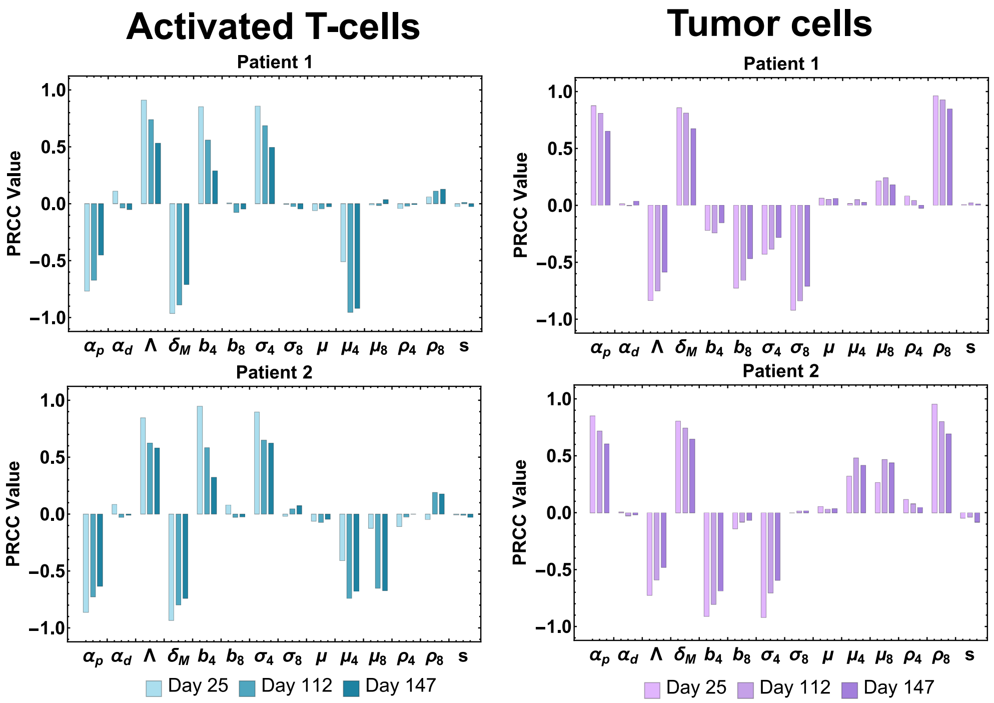


**Figure 3. Activated T-cells and Tumor cell population sensitivity.** PRCC values for the estimated parameters using the activated T-cells and tumor cell population as the output of interest.

#### B.9. Methods for the 'Application' section

Using all patient-specific parameters (Table 2 and parameters obtained from [1]), we created six different patient profiles, and varied their initial tumor cell counts, which are assumed to be contained in a spherical shaped tumor with diameter $d$ (see Table A5). To create a wide range of initial tumor cell counts corresponding to different tumor sizes (sample size: 2000), we used the LHS technique to get a uniform and equal distribution of tumor cell counts ranging from 30,000 to $2 \times 10^{10}$ cells. Notice that this set of different initial conditions implicitly correspond to a total number of cells $x$ number of days (or months) after having resection surgery. Keep in mind that there may be at least 5% tumor residue and may continue to grow post-surgery. Therefore, the estimated tumor cell count (or diameter) in Table A5 is the tumor cell count at day 0 of vaccine treatment.

**Table A5. Tumor cell count in a spherical shaped tumor with diameter $d$.**

| Tumor cell count range | Diameter $d$ (mm) |
|---|---|
| $(3 \times 10^4,\ 5 \times 10^4]$ | $\leq 1$ |
| $(5 \times 10^4,\ 5 \times 10^5]$ | $(1, 2.4]$ |
| $(5 \times 10^5,\ 1 \times 10^7]$ | $(2.4, 6.4]$ |
| $(1 \times 10^7,\ 5 \times 10^8]$ | $(6.4, 23.45]$ |
| $(5 \times 10^8,\ 2 \times 10^{10}]$ | $(23.45, 89.2]$ |

#### References


1. Ott PA, Hu Z, Keskin DB, Shukla SA, Sun J, Bozym DJ, et al. An immunogenic personal neoantigen vaccine for patients with melanoma. Nature. 2017;547(7662):217–221. doi:10.1038/nature22991.

2. Stothard P. The sequence manipulation suite: JavaScript programs for analyzing and formatting protein and DNA sequences.; 2000. Available from: `https://www.bioinformatics.org/sms/prot_mw.html`.

3. Classical and molecular biology;. Available from: `http://www.molbiol.ru/eng/scripts/01_04.html`.

4. Orsini G, Legitimo A, Failli A, Massei F, Biver P, Consolini R. Enumeration of human peripheral blood dendritic cells throughout the life. International immunology. 2012;24(6):347–356.

5. Walker J, Tough DF. Modification of TLR-induced activation of human dendritic cells by type I IFN: synergistic interaction with TLR4 but not TLR3 agonists. Eur J Immunol. 2006;36(7):1827–36. doi:10.1002/eji.200635854.

6. Mathematica, Version 12.0;. Available from: `https://www.wolfram.com/mathematica`.

7. Rohatgi A. Webplotdigitizer: Version 4.3; 2020. Available from: `https://automeris.io/WebPlotDigitizer`.

8. Li Z, Ju X, Silveira PA, Abadir E, Hsu WH, Hart DNJ, et al. CD83: Activation Marker for Antigen Presenting Cells and Its Therapeutic Potential. Frontiers in Immunology. 2019;10:1312. doi:10.3389/fimmu.2019.01312.

9. Stark PB. SticiGui, Onsophic, and Statistics W21. University of California, Berkeley. 2011;.

10. Bittersohl H, Steimer W. Intracellular concentrations of immunosuppressants. In: Personalized Immunosuppression in Transplantation. Elsevier; 2016. p. 199–226.

11. Perelson AS, Weisbuch G. Immunology for physicists. Reviews of modern physics. 1997;69(4):1219.

12. Smith JG, Liu X, Kaufhold RM, Clair J, Caulfield MJ. Development and validation of a gamma interferon ELISPOT assay for quantitation of cellular immune responses to varicella-zoster virus. Clinical and diagnostic laboratory immunology. 2001;8(5):871–879.

13. Wykes MN, Renia L. ELISPOT assay to measure peptide-specific IFN-$\gamma$ production. Bio-protocol. 2017;7(11):e2302–e2302.

14. Karulin AY, Caspell R, Dittrich M, Lehmann PV. Normal distribution of CD8+ T-cell-derived ELISPOT counts within replicates justifies the reliance on parametric statistics for identifying positive responses. Cells. 2015;4(1):96–111.

15. Liu W, Dowling JP, Murray WK, McArthur GA, Thompson JF, Wolfe R, et al. Rate of growth in melanomas: characteristics and associations of rapidly growing melanomas. Archives of dermatology. 2006;142(12):1551–1558.

16. Society AC. Melanoma Skin Cancer Stages; 2019. Available from: `https://www.cancer.org/cancer/melanoma-skin-cancer/detection-diagnosis-staging/melanoma-skin-cancer-stages.html`.

17. Seriesa AC, Narod S. Disappearing breast cancers. Current Oncology. 2012;19(2):59.

18. Agustin-Bunch F. How many cancer cells are in a tumor?;. Available from: `https://drfarrahcancercenter.com/how-many-cancer-cells-are-in-a-tumor/`.

19. Alliance MR. Understanding Melanoma Staging; 2020. Available from: `https://www.curemelanoma.org/about-melanoma/melanoma-staging/understanding-melanoma-staging/`.

20. Ganeshalingam S, Koh DM. Nodal staging. Cancer Imaging. 2009;9(1). doi:10.1102/1470-7330.2009.0017.

21. Shan Q, Fan Y, Guo J, Han X, Wang H, Wang Z. Relationship between tumor size and metastatic site in patients with stage IV non-small cell lung cancer: A large SEER-based study. PeerJ. 2019;7:e7822. doi:10.7717/peerj.7822.

22. Foote J, Eisen HN. Kinetic and affinity limits on antibodies produced during immune responses. Proceedings of the National Academy of Sciences of the United States of America. 1995;92(5):1254.

23. Marino S, Hogue IB, Ray CJ, Kirschner DE. A methodology for performing global uncertainty and sensitivity analysis in systems biology. J Theor Biol. 2008;254(1):178–96. doi:10.1016/j.jtbi.2008.04.011.
