## Supplementary material for "Mathematical Model of a Personalized Neoantigen Cancer Vaccine and the Human Immune System: Evaluation of Efficacy": S1 Table

### S1 Table. Parameter values and definition

**Table S1.** Parameter values and definition.(♣ estimated from literature; ♦ estimated from data (clinical/experimental), see S1 Appendix for details; ★ fitted to trial data)

| | Parameter | Definition | Value/Range | Value Used | Units |
|---|---|---|---|---|---|
| **Vaccine** | $Dose_p$ | Peptide concentration at each dose | clinical trial data [1] | patient-specific ♦ | pmol |
| | $Dose_a$ | Adjuvant concentration at each dose | | 500 [1, 2] | mg/L |
| | $\alpha_p$ | Internalization rate of peptides by DCs | | 0.28 [3] | $day^{-1}$ |
| | $\alpha_d$ | Internalization rate of adjuvant by DCs | | 0.5 | $day^{-1}$ |
| | $V_{sc}$ | Volume of injections site | - | $4 \times (0.1919)$ [4] | $L$ |
| **Dendritic cells** | $\Lambda$ | Maximum growth rate | | 3.75 | $day^{-1}$ |
| | $\delta_M$ | Death rate of mature DCs | (0.23,0.54) [5] | 0.33 | $day^{-1}$ |
| | $r_D$ | Maximum differentiation rate | | 2.48 ★ | $day^{-1}$ |
| | $K_a$ | Half-maximum adjuvant effect constant | | 6.64 ★ | - |
| | $K_{DC}$ | Carrying capacity | $(2.07, 2.68) \times 10^7$ ♦ | $2.38 \times 10^7$ (median) | cells |
| | $V_E$ | Volume of endosomes in a DC | $(0.16, 1.35) \times 10^{-14}$ [6] | $1 \times 10^{-14}$ | $L$ |
| **Antigen processing and presentation** | $N_A$ | Avogadro's constant | - | $6.02 \times 10^{23}$ | $mol^{-1}$ |
| | $\alpha_p^E$ | Endosomal internalization rate of peptides | [10.8,144] [6] | 70 | $day^{-1}$ |
| | $\beta_p$ | Degradation rate of peptides | 14.4-21.6 [7] | 14.4 | $day^{-1}$ |
| | $k_{\text{on},1}$ | On rate for T-epitope-MHCI binding with allele $j$ | | $1.8144 \times 10^{-2}$ [8] | $pM^{-1} \cdot day^{-1}$ |
| | $k_{\text{off},j}$ | Off rate for T-epitope-MHCI binding with allele $j$ | Neoantigen specific (based on $K_{D,j}^{\text{eff}}$) | $k_{\text{on},1} \cdot K_{D,j}^{\text{eff}}$ | $day^{-1}$ |
| | $k_{\text{on},2}$ | On rate for T-epitope-MHCII binding with allele $k$ | $[0.43, 4.3] \times 10^{-2}$ [6, 9] | $8.64 \times 10^{-3}$ ♦ | $pM^{-1} \cdot day^{-1}$ |
| | $k_{\text{off},k}$ | Off rate for T-epitope-MHCII binding with allele $k$ | Neoantigen specific (based on $K_{D,k}^{\text{eff}}$) | $k_{\text{on},2} \cdot K_{D,k}^{\text{eff}}$ | $day^{-1}$ |
| | $\beta_M$ | Degradation rate of endosomal free MHCI/II molecules | 0.92-2.08 [6] | 1.663 [6] | $day^{-1}$ |
| | $k_{\text{in}}$ | Recycling rate of free MHC molecules | 10.8-14.4 [6, 7] | 14.4 [6] | $day^{-1}$ |
| | $k_{\text{ext}}$ | Exocytosis rate of p-MHC complex | 17.28-43.2 [6, 7] | 28.8 [6] | $day^{-1}$ |
| | $\beta_{pM}$ | Degradation rate of p-MHCI/II complex | $\geq 0.1663$ [6] | 0.166 [6] | $day^{-1}$ |

Continued on next page

| | Parameter | Description | Value/Range | Value Used | Units |
|---|---|---|---|---|---|
| **T-cells** | $a$ | Number of tumor cells needed for half-maximal $A_{CD8}$ proliferation | | $5 \times 10^7$ [10] | cells |
| | $a_1$ | Half saturation constant of the CD4$^+$ T-cells production rate | | $1 \times 10^3$ [10–12] | cells |
| | $b_4$ | Max. growth rate of naïve CD4$^+$ T-cells | | 0.15 [13] | day$^{-1}$ |
| | $b_8$ | Max. growth rate of naïve CD8$^+$ T-cells | | 0.12 [13] | day$^{-1}$ |
| | $\sigma_4$ | Max. activation rate of naïve CD4$^+$ T-cells | | 1.5 [6, 14] | day$^{-1}$ |
| | $\sigma_8$ | Max. activation rate of naïve CD8$^+$ T-cells | | 3 [14] | day$^{-1}$ |
| | $K_{TC}$ | Carrying capacity of T-cells | | $8.57 \times 10^{11}$ ♣ | cells |
| | $F_{P_4}$ | Frequency of antigen-specific CD4$^+$ T-cells | (0,0.01) [1] | $\{0.006, 0.001, 0.001, 0.001, 0.003, 0.001\}$ | - |
| | $F_{P_8}$ | Freq. of antigen-specific CD8$^+$ T-cells | (0,0.01) [1] | $\{0.007, 0.002, 0.006, 0.002, 0.002, 0.001\}$ | - |
| | $K_{pM}$ | Half-maximum effect of activation | 400 [3, 6] | 400 | - |
| | $c$ | Maximum CD8$^+$ T-cells recruitment rate | | patient-specific★ | day$^{-1}$ |
| | $c_4$ | Maximum CD4$^+$ T-cell production rate | | patient-specific★ | day$^{-1}$ |
| | $c_8$ | Rate at which CD8$^+$ T-cells are stimulated to be produced | | $6.5 \times 10^{-11}$ [12, 15, 16] | day$^{-1}$ |
| | $\mu$ | Death rate of naïve T-cells | (0.00014,0.012) [13, 17] | 0.0029 | day$^{-1}$ |
| | $\mu_4$ | Death rate of activated CD4$^+$ T-cells | (0.022,0.083) [18] | 0.031 | day$^{-1}$ |
| | $\mu_8$ | Death rate of activated CD8$^+$ T-cells | (0.022,0.052) [18] | 0.022 | day$^{-1}$ |
| | $\rho_4$ | Proliferation rate for activated T-cells | (0.0053,0.0477) [13] | 0.0265 | day$^{-1}$ |
| | $\rho_8$ | Proliferation rate for activated CD8$^+$ T-cells | (0,0.1106) [13] | 0.0509 | day$^{-1}$ |
| **Tumor cells** | $r$ | Maximum growth rate | tumor-type dependent | 0.004 ♣ | day$^{-1}$ |
| | $K_T$ | Carrying capacity of tumor | | $1.45 \times 10^{10}$ ⧫ | cells |
| | $d$ | Max. lysis rate by activated T-cells | (0.01,0.05) [19] | patient-specific★ | day$^{-1}$ |
| | $\lambda$ | Dependence of lysis rate on the effector/target ratio constant | (0,1) | patient-specific★ | - |
| | $s$ | Half-maximal effect of tumor cell lysis | | 0.0839 [12] | - |

Continued on next page

**Table S1 – continued from previous page**

| | Parameter | Description | Value/Range | Value Used | Units |
|---|---|---|---|---|---|
| **Initial Conditions** | $A_d(0)$ | Adjuvant concentration in a vaccine | | 500 | mg/L |
| | $p(0)$ | Peptide amount in a vaccine | | patient-specific ⧫ | pmol |
| | $D_I(0)$ | Immature dendritic cells | | $1 \times 10^7$ | cells |
| | $D_M(0)$ | Mature dendritic cells | | 0 | cells |
| | $p^E(0)$ | Endosomal peptides | | 0 | pmol |
| | $M_j^E(0)$ | Endosomal MHC-I | | $(1.6, 5.8) \times 10^{-7}$ [3,6] | pmol |
| | $M_k^E(0)$ | Endosomal MHC-II | | $(0.16, 8.3) \times 10^{-8}$ [3,6] | pmol |
| | $pM_j^E(0)$ | Endosomal p-MHCI complex | | 0 | pmol |
| | $pM_k^E(0)$ | Endosomal p-MHCII complex | | 0 | pmol |
| | $pM_j(0)$ | p-MHCI on DC membrane | | 0 | pmol |
| | $pM_k(0)$ | p-MHCII on DC membrane | | 0 | pmol |
| | $M_j(0)$ | Free MHC-I on DC membrane | | 0 | pmol |
| | $M_k(0)$ | Free MHC-II on DC membrane | | 0 | pmol |
| | $N_{CD4}(0)$ | Naïve $CD4^+$ T-cell count | $(2.15, 8.6) \times 10^9$⧫ | $(5.38 \times 10^9) \times 0.7$ | cells |
| | $N_{CD8}(0)$ | Naïve $CD8^+$ T-cell count | $(2.15, 8.6) \times 10^9$⧫ | $(5.38 \times 10^9) \times 0.3$ | cells |
| | $A_{CD4}(0)$ | Activated $CD4^+$ T-cell count | patient-specific ⧫ | | cells |
| | $A_{CD8}(0)$ | Activated $CD8^+$ T-cell count | patient-specific ⧫ | | cells |
| | $T(0)$ | Tumor cell count | | patient-specific ⧫ | cells |

#### References


1. Ott PA, Hu Z, Keskin DB, Shukla SA, Sun J, Bozym DJ, et al. An immunogenic personal neoantigen vaccine for patients with melanoma. Nature. 2017;547(7662):217–221. doi:10.1038/nature22991.

2. Keskin DB, Anandappa AJ, Sun J, Tirosh I, Mathewson ND, Li S, et al. Neoantigen vaccine generates intratumoral T cell responses in phase Ib glioblastoma trial. Nature. 2019;565(7738):234–239. doi:10.1038/s41586-018-0792-9.

3. Chen X, Hickling T, Vicini P. A mechanistic, multiscale mathematical model of immunogenicity for therapeutic proteins: part 1—theoretical model. CPT: pharmacometrics & systems pharmacology. 2014;3(9):1–9.

4. Tegenge MA, Mitkus RJ. A physiologically-based pharmacokinetic (PBPK) model of squalene-containing adjuvant in human vaccines. Journal of pharmacokinetics and pharmacodynamics. 2013;40(5):545–556.

5. Lanzavecchia A, Sallusto F. Regulation of T cell immunity by dendritic cells. Cell. 2001;106(3):263–266.

6. Yogurtcu ON, Sauna ZE, McGill JR, Tegenge MA, Yang H. TCPro: an In Silico Risk Assessment Tool for Biotherapeutic Protein Immunogenicity. The AAPS journal. 2019;21(5):96.

7. Singer DF, Linderman JJ. The relationship between antigen concentration, antigen internalization, and antigenic complexes: modeling insights into antigen processing and presentation. The Journal of cell biology. 1990;111(1):55–68.

8. Corr M, Slanetz A, Boyd L, Jelonek M, Khilko S, al Ramadi B, et al. T cell receptor-MHC class I peptide interactions: affinity, kinetics, and specificity. Science. 1994;265(5174):946–949. doi:10.1126/science.8052850.

9. Kasson PM, Rabinowitz JD, Schmitt L, Davis MM, McConnell HM. Kinetics of peptide binding to the class II MHC protein I- Ek. Biochemistry. 2000;39(5):1048–1058.

10. Hu X, Jang SRJ. Dynamics of tumor–CD4+–cytokine–host cells interactions with treatments. Applied Mathematics and Computation. 2018;321:700–720.

11. Arciero J, Jackson T, Kirschner D. A mathematical model of tumor-immune evasion and siRNA treatment. Discrete & Continuous Dynamical Systems-B. 2004;4(1):39.

12. Makhlouf AM, El-Shennawy L, Elkaranshawy HA. Mathematical modelling for the role of CD4+ T cells in tumor-immune interactions. Computational and mathematical methods in medicine. 2020;2020.

13. Macallan DC, Asquith B, Irvine AJ, Wallace DL, Worth A, Ghattas H, et al. Measurement and modeling of human T cell kinetics. European journal of immunology. 2003;33(8):2316–2326.

14. Lee HY, Topham DJ, Park SY, Hollenbaugh J, Treanor J, Mosmann TR, et al. Simulation and prediction of the adaptive immune response to influenza A virus infection. Journal of virology. 2009;83(14):7151–7165.

15. de Pillis LG, Radunskaya AE. Modeling tumor–immune dynamics. In: Mathematical Models of Tumor-Immune System Dynamics. Springer; 2014. p. 59–108.

16. de Pillis LG, Gu W, Radunskaya AE. Mixed immunotherapy and chemotherapy of tumors: modeling, applications and biological interpretations. J Theor Biol. 2006;238(4):841–62. doi:10.1016/j.jtbi.2005.06.037.

17. Mclean AR, Michie CA. In vivo estimates of division and death rates of human T lymphocytes. Proceedings of the National Academy of Sciences. 1995;92(9):3707–3711.

18. Ribeiro RM, Mohri H, Ho DD, Perelson AS. In vivo dynamics of T cell activation, proliferation, and death in HIV-1 infection: why are CD4+ but not CD8+ T cells depleted? Proceedings of the National Academy of Sciences. 2002;99(24):15572–15577.

19. Rhodes A, Hillen T. A mathematical model for the immune-mediated theory of metastasis. Journal of theoretical biology. 2019;482:109999.
